# The omnicausal model reveals the highly polyfactorial nature of complex diseases

**DOI:** 10.1101/2025.08.20.25333988

**Authors:** Carla Márquez-Luna, Martin Tournaire, Ghislain Rocheleau, Ron Do, Marie Verbanck

**Affiliations:** The Charles Bronfman Institute for Personalized Medicine, Icahn School of Medicine at Mount Sinai, New York, New York, USA; Department of Genetics and Genomic Sciences, Icahn School of Medicine at Mount Sinai, New York, New York, USA; UR 7537 BioSTM, Biostatistique, Traitement et Modélisation des données biologiques, Université Paris Cité, France; Institut Curie, PSL Research University, Inserm U1331 Computational Oncology, Team Genetic Epidemiology of Cancers, 75248 Paris, France

**Keywords:** Mendelian randomization, complex diseases, phenome-wide risk factors, causality explained, polyfactorial index, omnicausal model

## Abstract

Mendelian Randomization (MR) is a human genetics method for inferring causal relationships between risk factors and diseases. A common focus of MR studies has been on the causal inference of a single risk factor on a single disease. This has led to the successful discovery of numerous causal risk factors for disease. However, it remains unclear how much each causal risk factor contributes to disease collectively. Here, we introduce the concept of “causality explained”, that provides an estimate of the causal variance explained by a phenome-wide set of risk factors on complex diseases to assess how much causality can be potentially explained. The model is based on principal component regression which is a multivariate linear regression based on principal component analysis. In complement, we propose the “polyfactorial index” to assess the trajectory of causality explained as risk factors are sequentially added into the model, to characterize the causal architecture for a complex disease. We demonstrate that our model correctly assesses the causality explained and causal architecture in simulations across a wide range of parameters. To build our model, we used a phenome-wide set of 222 traits from the UK Biobank compared to a set of 5 known risk factors for coronary artery disease. We observed that the phenome-wide set explains almot 45% of causality compared to 28.73% for the set of known risk factors. In addition, we tested our approach on 13 complex diseases and showed that the phenome-wide set can explain between 27% for anorexia to 80% for schizophrenia, with increasing trajectories of causality explained. We propose the “omnicausal model”, which posits that a large number of risk factors explains a very small portion individually to disease but collectively explain most of the causal variance. We distinguished core and peripheral causal factors that explain respectively a larger and a smaller part of causal variance. This approach provides insights into the relative importance of individual risk factors as well as the collective impact of multiple causal risk factors on disease, providing insights into the causal architecture of disease.

## Introduction

Mendelian Randomization (MR) is a human genetics approach that allows for causal inference of risk factors on disease outcome^1^. MR is analogous to randomized controlled trials where the drug and placebo groups are substituted by genotype groups of a genetic variant that is naturally associated with differential levels of the exposure trait (e.g. low-density lipoprotein-cholesterol) to estimate its causal effect on the outcome trait (e.g. coronary artery disease). Hence, MR can overcome some of the limitations of observational studies including reverse causation, confounding, and additional biases^2^. In two-sample MR, genetic variant summary statistics, i.e. the estimated effect sizes and standard errors, are retrieved from genome-wide association studies (GWASs) on the exposure and outcome traits. Then, a linear regression can be performed between the outcome effect sizes and the exposure effects across multiple exposure-associated genetic variants. The slope of the regression thus provides an estimator of the causal effect of the exposure on the outcome. However, MR is sensitive to three crucial assumptions related to the absence of pleiotropy: 1) the genetic variants, called instrumental variables (IVs), must be strongly associated with the exposure; 2) the genetic variants are independent of confounding factors of the exposure-outcome relationship, i.e. no correlated pleiotropy; and 3) the genetic variants are only associated with the disease outcome through the risk factor of interest, i.e. no horizontal pleiotropy. Studies have shown that pleiotropy is highly prevalent in human genetic variation^3^. Moreover, human complex traits display an omnigenic architecture^4^, *i*.*e*. are influenced by an infinitely large number of genes with small individual contributions, emphasizing the pervasive nature of pleiotropy. In a previous study, we detected horizontal pleiotropy in more than ∼50% of inferred causal relationships from 4,250 Mendelian randomization tests. We showed that horizontal pleiotropy can bias causal estimates in MR testing and result in false-positive discoveries^5^.

Several MR studies have been conducted, leading to the identification of many causal risk factors for various diseases^6^. These studies have typically focused on testing the causality of a single risk factor on a single disease, although recent studies have extended MR to testing a single risk factor across a large number of diseases in a phenome-wide manner^7^. In addition, detecting and correcting for pleiotropy in MR has been an active field of research and has led to the development of many novel statistical methods^5,8–13^. Nevertheless, not a single method is effective for all types of pleiotropy^6^, and therefore, most current methods, which are based on single-variable MR (SVMR), presumably suffer from massive pleiotropy. With a different strategy, it has been shown that pleiotropy can be taken into account in MR if the second exposure responsible for the pleiotropic effect of the variant is included in the MR model^14,15^ into a multi-variable Mendelian Randomization (MVMR) model.

Although much progress has been made in identifying causal risk factors for complex diseases, not much is known about the overall causal contribution that a particular risk factor has on disease. Quantifying the overall causal contribution can be useful in at least three ways: 1) to understand the relative importance of a single causal risk factor to another; 2) to evaluate the contribution of multiple risk factors collectively on a single disease; and 3) to elucidate the causal architecture of risk factors for a complex disease. Here, we introduce the concept of “causality explained”, which represents the variance explained by one or several causal risk factors on disease in a MR model. In addition, in order to study the causality explained trajectory when adding risk factors one by one, we propose the “polyfactorial index”. Both metrics allow us to characterize the causal architecture of risk factors for a complex disease. After validating our model on simulations, we applied our method to the study of coronary artery disease using a phenome-wide set of exposures from the UK Biobank and compared it with the causality explained by well-known risk factors. Additionally, we applied our model to a range of complex diseases using the same set of exposures from the UK Biobank. Furthermore, we propose the “omnicausal model” which posits that a large number of risk factors explain a very small portion of the causality explained individually but a large portion collectively, resulting in a causality explained trajectory that continually increases as risk factors are added.

## Results

### Overview of the methodology

We introduce the concept of “causality explained” as the portion of variance attributed to one or more risk factors in the context of Mendelian Randomization (MR). The causality explained can be assessed in the context of single-variable Mendelian randomization (SVMR) in a MR model testing the effect of one exposure on one disease outcome. However, the concept of causality explained can also be applied in the context of multivariable (MVMR) where multiple exposures are tested for causality on a disease outcome. In MVMR, all exposures with at least one instrumental variable (IV) (*i*.*e*. one independent genome-wide-significant genetic variant) are combined into a multivariate linear regression model, all the exposure-associated IVs being considered as the observations in the model. In addition, in the causality explained MR model, we propose to use an exhaustive set of potential risk factors, such as phenome-wide exposures, to quantify the amount of causality that these risk factors are expected to explain altogether. Therefore, principal component analysis is used to circumvent issues arising from colinearities between exposures, and the multivariate linear regression model is run on a subset of significant principal components. The amount of causality explained is then measured by the *R*^2^, which is the proportion of variation, representing causality, in the disease outcome that is predictable from all the exposures altogether. The *R*^2^ is adjusted for the number of exposures in the model. Additionally, the prediction errors from the model used in the *R*^2^ correspond to cross-validated prediction errors and are calculated from a leave-one-out procedure, sequentially omitting each IV to compute prediction errors.

In addition, we propose to study the trajectory of causality explained as exposures are sequentially incorporated into the model, ordered by their individual level of causality explained. We introduce a complementary metric called the “polyfactorial index” that allows to characterize the trajectory of causality explained. The polyfactorial index represents the maximum difference in causality explained between the observed and theoretical trajectories of causality explained, with the theoretical trajectory corresponding to a monotonous increase from the percentage of causality explained by the first exposure to 100%. In theory, the maximum value of the polyfactorial index would be close to 1, and a high polyfactorial index mean that the causality explained increases very rapidly with only a few exposures. On the contrary, the mininum value of the polyfactorial index is zero (negative values are set to zero), and means that the causality explained increases very slowly and each added exposure contributes to the maximum level of causality explained.

Interpreted jointly, the causality explained and the polyfactorial index allow to characterize the causal architecture of a complex disease in terms of risk factors.

### Evaluation of the statistical properties of the causality explained model in simulations

We evaluated our causality explained model and metrics on simulations where we simulated a disease outcome with both causal and non-causal exposures. Specifically, we simulated a disease outcome *Y* with a network of causality constituted of 4 distinct subsets of exposures denoted *A, B, C*, and *D*, with the total number of potential exposures equal to 100 (See Methods and Figure 4 for details). Only exposures from subsets *A* and *C* were causal to the disease outcome *Y*, whereas exposures from *B* and *D* were not causal to *Y*. In addition, exposures from *A* were causal to exposures from *B*, inducing vertical pleiotropy. Finally, exposures from *D* were causally linked to neither any other exposure nor the disease outcome. We varied the number of exposures from *A* and *C* that were causal to the disease outcome *Y*. Since the total number of potential exposures (from all four sets) was fixed to 100, when the number of causal exposures from *A* and *C* increases, the number of non-causal exposures from *B* and *C* decreases accordingly. Scenarios included either 4, 20, 40, 60, 80, or 100 causal exposures out of a 100 total exposures. Each exposure was simulated with a certain proportion of causal genetic variants that can be used as instrumental variables (IVs) in the MR model. In addition, the pleiotropic architecture of the exposures was modified by varying the proportion of variants with pleiotropic effects on at least two causal exposures from *A* and *C* inducing horizontal pleiotropy. Finally, a confounder *U* was simulated between each exposure and outcome, inducing correlated pleiotropy. We ran our causality explained model by first identifying IVs for all the exposures (*i*.*e* variant with an association p-value < 5 × 10^-8^) and ordering all exposures according to decreasing individual causality explained. Second, to compute the causality explained trajectory, we run a MVMR model with principal component analysis by sequentially including the previously ordered exposures in the model (See Methods for details).

First, we studied the trajectory of the causality explained according to the size of the model, corresponding to the sequential incorporation of exposures from 1 to 100 (Figure 1A). We observed that the percentage of causality explained increased steadily until reaching a plateau as exposures were incorporated into the model. Interestingly, the causality explained trajectory showed an inflection point precisely when the number of included exposures equaled the true number of causal exposures (in the different panes).

**Figure 1.**
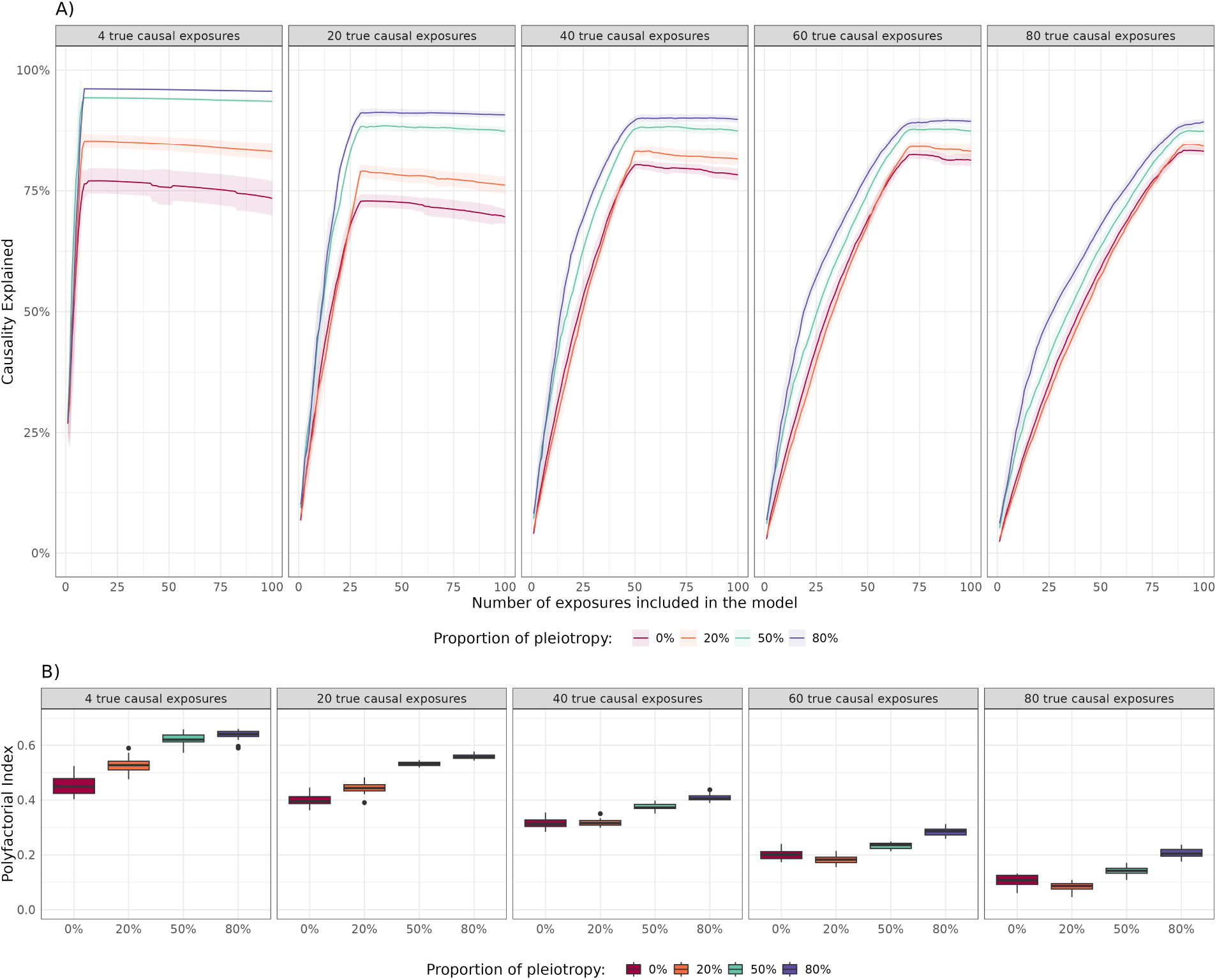
Evaluation of the causality explained model in simulation. In panel **A)**, the y-axis represents the evolution of the average causality explained and standard error over 20 replicates, according to the size of the multivariable Mendelian randomization model (x-axis), by sequentially including exposures into the model. At each step principal component analysis is used to account for colinearities. In panel **B)**, the y-axis represents the polyfactorial index that characterizes the trajectory of causality explained when adding exposures one by one according to the causal variants pleiotropy (x-axis). In both panels, the panes distinguish the true number of causal exposures, and colors represent different pleiotropic architectures induced by the proportion of causal variants with pleiotropic effect on at least two exposures.

It is worth noting that figure 1A shows the results averaged over 20 replicates but the trajectories were not that smooth when looking at the replicates one by one without, even if the interpretation remained similar (Supplementary Figure 1). Importantly, the maximum causality explained did not reach 100% even when all 100 potential exposures were included in the model. This was partly due to the noise included in the model, and mostly because only a subset of IVs were detected and included in the model. Indeed, in our simulations, we first ran a GWAS to identify instrumental variables (IVs) for each exposure and did have 100% power to detect these IVs. When considering the scenarios without pleiotropy, we could see that the maximum causality explained tended to be closer to 100% when the number of true causal exposures increased, due to an increase in the total pool of detected IVs whatever the exposure.

To characterize the trajectory, we calculated the polyfactorial index (Figure 1B) that was able to distinguish between the different causal architectures of outcomes. Indeed the polyfactorial index increased as the number of true causal exposures increased. Therefore, the lower the polyfactorial index, the more polyfactorial the outcome. In simulations, the polyfactorial index ranged from 0.028 to 0.662. The higher the polyfactorial index, the closer to the monofactorial outcome, whereas a polyfactorial index close to 0 indicates a highly polyfactorial outcome for which the causality explained kept increasing as more exposures were added into the model. In addition, as illustrated in Supplementary Figure 2, when the per-variant heritability decreased from 0.008 to 0.004, both the maximum causality explained and the polyfactorial index only slightly decreased due to a decrease in the power to detect IVs.

Pleiotropy seemed to have a major effect on both the maximum causality explained and the polyfactorial index. Indeed, the proportion of pleiotropic variants increased the causality explained especially when causal exposures were missing from the model, that is before reaching the plateau corresponding to all true causal exposures included in the model. Indeed, in the case where genetic variants have pleiotropic effects on at least two causal exposures, these variants can serve as IVs for multiple exposures, which allows to explain a higher proportion of causality even when causal exposures are missing from the model (Supplementary Figure 3). As a consequence, the polyfactorial index also increased with the proportion of pleiotropy. This is a strong feature of our causality explained model since pleiotropy is widespread in the human genome^3,16,17^, we expect to capture a larger proportion of causality even if we are missing some causal exposures.

In addition, we assessed how the causal relationship structure affected the causality explained by looking at the impact of vertical pleiotropy for non-causal exposures. Specifically, we simulated 50 causal exposures (10 from set *A* and 40 from set *C*), and 50 non-causal exposures. Then, we varied the proportion of non-causal exposures coming from *B* with a vertical pleiotropic effect of exposures from *A* (*i*.*e*. exposure with a causal effect from exposures in *A*), and from *C* without any vertical pleiotropic effect. We observed that even if none of the exposures from *B* were causal to the outcome, increasing the number of non-causal exposures involved in vertical pleiotropy effects with causal exposures increased the causality explained (Supplementary Figure 4). Again, this is advantageous for the causality explained model since we do expect real exposures to be submitted to complex patterns of vertical pleiotropy.

On a different note, we defined the causality gain as the difference between the causality explained by the first exposure and the maximum causality explained. As illustrated in Figure 1, in simulations the causality explained by the first exposure included in the model varied according to the total number of true causal exposures. Indeed in the case of a highly polyfactorial outcome, the first exposure explains a lower proportion of causality explained, and therefore the causality gain is higher (Supplementary Figure 5). Moreover, both the causality gain and the polyfactorial index depended on the proportion of pleiotropy, particularly in lower polyfactorial outcomes. Therefore it is important to study the causality explained trajectory in light of the causality gain and the polyfactorial index.

Finally, to account for colinearities we used a principal component (PC) analysis, and we checked that the PC selection procedure did not affect the interpretation of the causality explained trajectory and polyfactorial index. We used an alternative strategy to select the PCs which consisted in retaining PCs with eigenvalues greater than 1 in the standardized PC analysis (See Methods for details). As illustrated in Supplementary Figure 6, the results were highly similar to Figure 1.

### Using all PheCodes explains almost half of the causality for coronary artery disease (CAD)

Coronary artery disease (CAD) is a heritable complex disease that is one of the leading causes of death worldwide. Many genetic variants have been identified as associated with CAD^18^, but collectively explain a small proportion of heritability which is estimated between 40 and 60% from twin studies^19^. MR studies have identified multiple risk factors that are causal for CAD, such as low-density lipoproteins (LDL) cholesterol and triglycerides^14^. However, CAD is a heterogeneous disease influenced by multiple pathways that implicate multiple related risk factors and comorbidities. Therefore, to better understand the etiology of CAD, it is important 1) to assess the relative importance of these established causal risk factors, and 2) to quantify the amount of causality expected to be explained by an exhaustive set of risk factors. For example, it is unknown how much each of the established causal risk factors for CAD - including LDL cholesterol, plasma triglycerides, blood pressure, body mass index, type 2 diabetes, and lipoprotein(a) - explain collectively on CAD and how much remains to be explained. Therefore, we chose to apply the causality-explained model to CAD.

As we have shown in simulations, the most exhaustive set of exposures explains the largest amount of causal variance. Hence, we aimed to include a large number of traits and diseases in our causality-explained model, leveraging phenome-wide association study (PheWAS) summary statistics based on a comprehensive set of PheCodes from the UK Biobank^20^, which are derived from electronical medical records, and span a broad range of medical diagnoses and conditions. Summary statistics for coronary artery disease (CAD) were retrieved from the CARDIoGRAMplusC4D Consortium^21^. Using these PheWAS summary statistics as exposures and CARDIoGRAMplusC4D summary statistics as the outcome, we quantified the causality explained by 1) known risk factors; and 2) an exhaustive set of traits on CAD. First, we performed single-variable MR (SVMR) on each of the known risk factors for CAD to estimate the causality explained for each factor separately. The known risk factors that we included were PheCodes analogous to the traditional risk factors. Specifically, we used PheCodes representing cholesterol increase: 272 - Disorders of lipoid metabolism, 272.1 - Hyperlipidemia and 272.11: Hypercholesterolemia; as well as diabetes: 250.2 - Type 2 diabetes; and finally blood pressure: 401 - Hypertension. After running the SVMR model for each one of these PheCodes, we observed that the largest individual contributions were that for Hyperlipidemia (28.53%), Disorders of lipoid metabolism (28.44%), Hypercholesterolemia (28.11%), Hypertension (11.88%), and Type 2 diabetes (5.04%). We next evaluated a combined MR model using MVMR to estimate the causality explained by all of these known risk factors altogether. We observed that the known risk factors for CAD collectively explain 28.73% of the causal variance. Second, we evaluated the contribution of an exhaustive set of PheCodes to the causality explained. To this aim, we included 218 PheCodes (with at least 1 IVs) collectively after filtering out Phecodes redundant with CAD (See Methods and Supplementary Table 2). We observed that this full set of 218 PheCodes explains almost 45% of the causality for CAD, which is more than twice the causality explained by known risk factors altogether (Table 1). It is worth mentioning that results on the summary statistics for CAD from the CARDIoGRAM consortium were highly similar with slightly lower levels of causality explained (Supplementary Table 1).

**Table 1.**
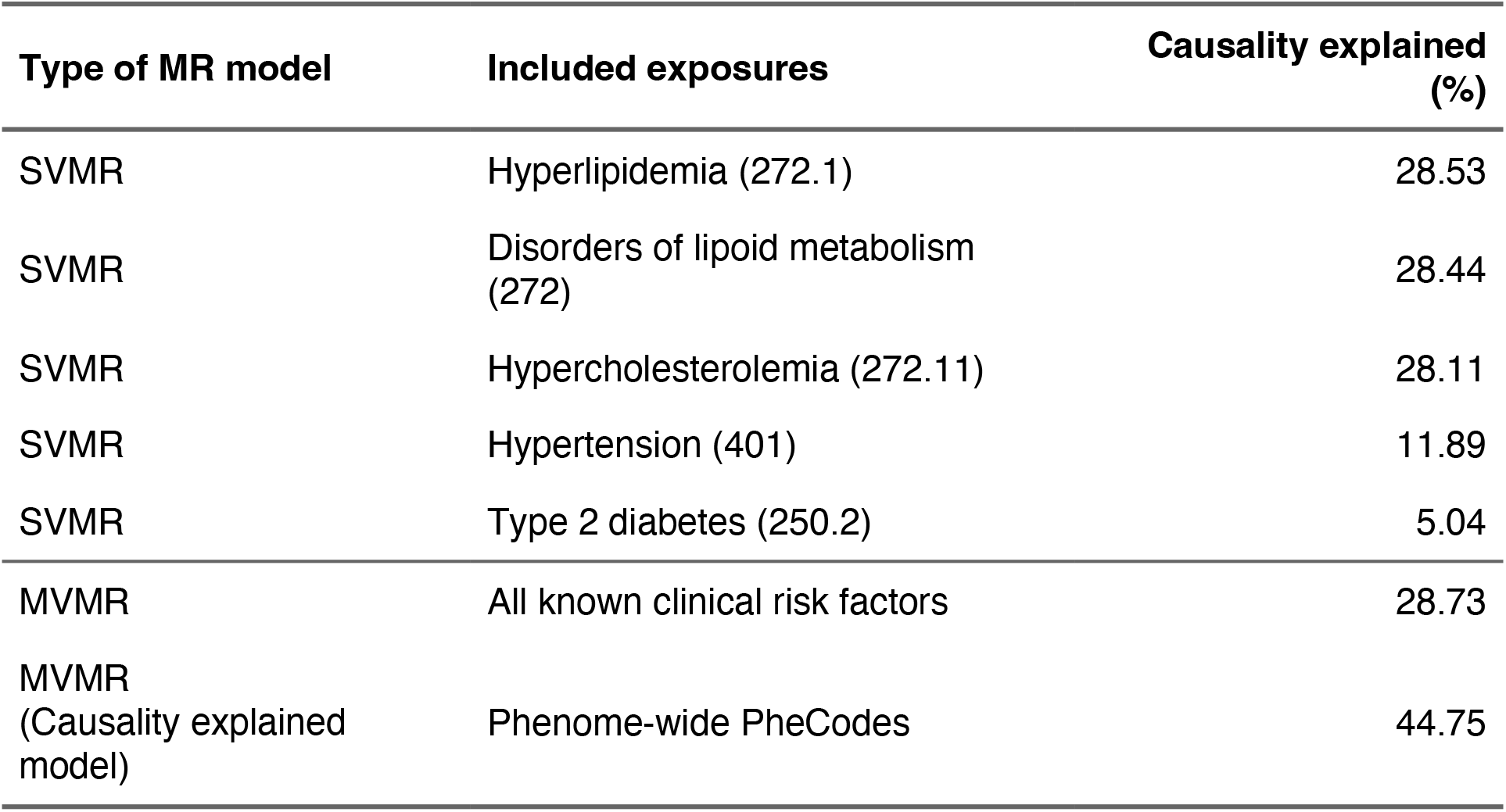
Causality explained in coronary artery disease (CAD) in a single-variable Mendelian randomization (SVMR) model for known risk factors individually, or in a multi-variable Mendelian randomization (MVMR) model for all known risk factors or phenome-wide set of PheCodes.

### The causality explained model distinguishes different causal architectures for 13 complex diseases

Next, we sought to extend the study of the causality explained in a wide range of heritable complex diseases. We included metabolic disorders: type-2 diabetes and chronic kidney disease; inflammatory diseases: Crohn’s disease, inflammatory bowel disease, rheumatoid arthritis, and ulcerative colitis; and finally psychiatric disorders: anorexia nervosa, autism spectrum disorder, bipolar disorder, major depressive disorder, psychiatric cross-disorder, and schizophrenia. CAD was also included for comparison purposes, making it a total of 13 complex diseases. We selected this set of 13 diseases with an understanding that these diseases have different genetic architectures, and therefore we expect different trajectories of causality explained and hence different causal architectures. For each one of these 13 diseases, we retrieved summary statistics from the corresponding GWAS analysis and ran our causality explained model using the set of maximum 222 PheCodes from the PheWAS on the UK Biobank after excluding disease-specific redundant PheCodes (*i*.*e*. between 186 and 221 included PheCodes, See Methods and Supplementary Table 2).

We compared the trajectories of causality explained for all 13 diseases (Figure 2), and what is particularly striking is that the causality explained kept increasing for all the diseases as more PheCodes were sequentially increased in the model. In addition, we could observe that the trajectories were quite different between the three categories of diseases, with a tendency for a steadier increase for psychiatric diseases than inflammatory diseases, as well as differences in maximum causality explained. Since the set of PheCodes was slightly different for each disease according to the exclusion of redundant PheCodes, we checked that the trajectories were similar when restricting to a set of common PheCodes (Supplementary Figure 7). Importantly, we wished to verify that the increasing causality explained trajectories that we could observe for all diseases was not an artifact. For a given size of the causality explained model, we permuted the effect sizes of all subsequent PheCodes included in the model (Supplementary Figure 8). Reassuringly, when the subsequently added Phecodes were permuted, the causality explained reached a plateau and even slightly decreased as expected.

**Figure 2.**
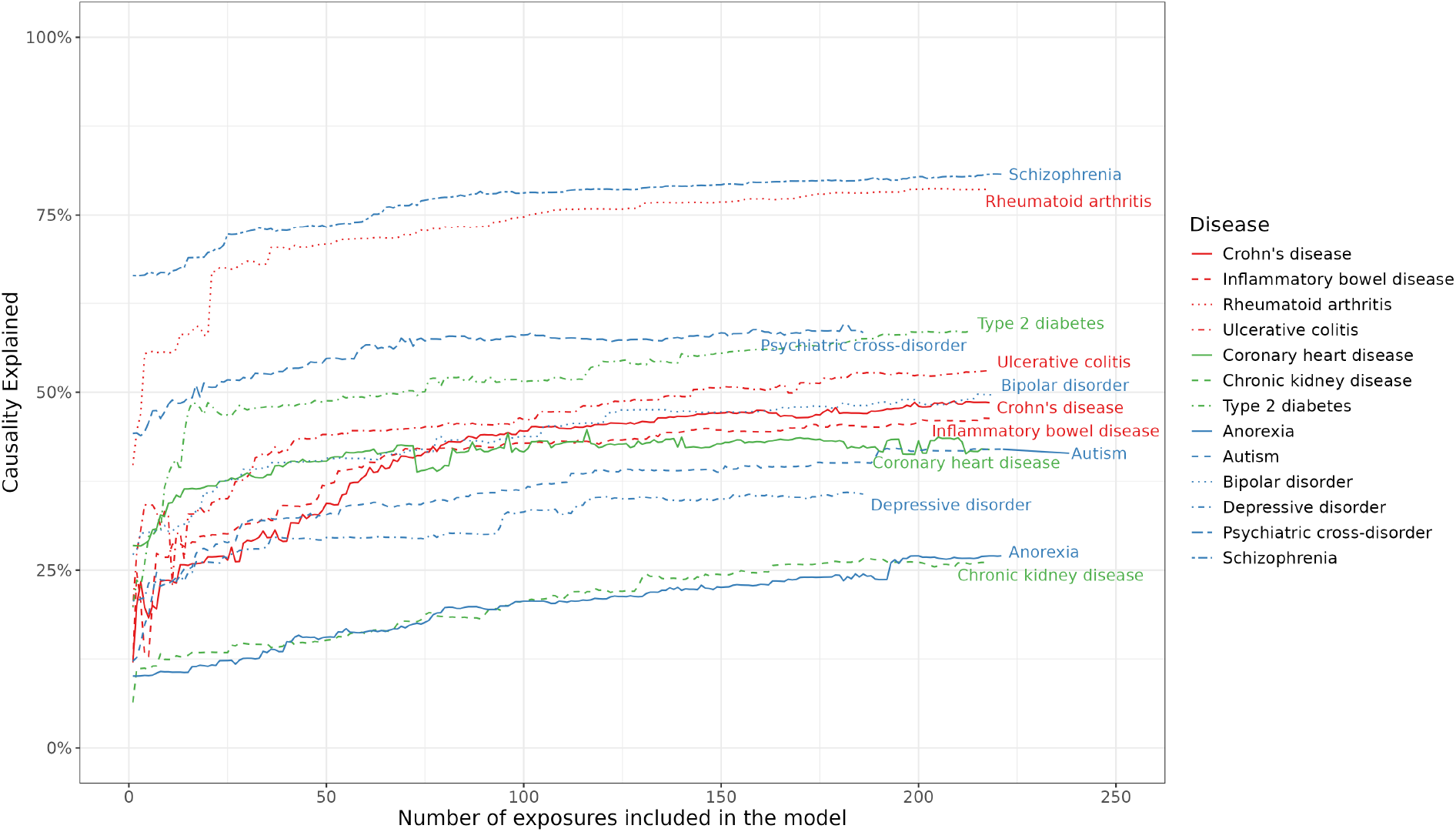
Evolution of the causality explained (y-axis) in 13 complex diseases according to the size of the multivariable Mendelian randomization model (x-axis) by sequentially including PheCodes from the UK Biobank, ordered by increasing individual causality explained, into the model. Diseases are color-coded according to broad categories.

In addition to the causality explained trajectories, we studied the associated metrics. We could observe that, although the maximum causality explained could be very different for complex diseases in the same category, the polyfactorial index seemed to be very close for related diseases (Figure 3). Indeed, the polyfactorial index was very low, ranging from 0 to 0.030 for 5 of the psychiatric traits (anorexia, bipolar disorder, depressive disorder, psychiatric cross-disorder, and schizophrenia). Similarly, the polyfactorial index was similar for Crohn’s disease, inflammatory bowel disease, and ulcerative colitis between 0.106 ans 0.133. This seemed to make sense because we can expect related complex diseases to have similar causal architectures and to be caused by similar risk factors. However, the maximum causality explained was very different, especially for psychiatric traits where it goes from 27% for anorexia to 80% for schizophrenia. Another interesting example is type 2 diabetes for which known risk factors have been identified such as weight, fat distribution or lipid levels for instance and yet this disease presents the highest polyfactorial index (0.23). This means that although major causal risk factors explain a certain amount of causality, using a phenome-wide set of exposures allows to significantly increase the causality explained.

**Figure 3.**
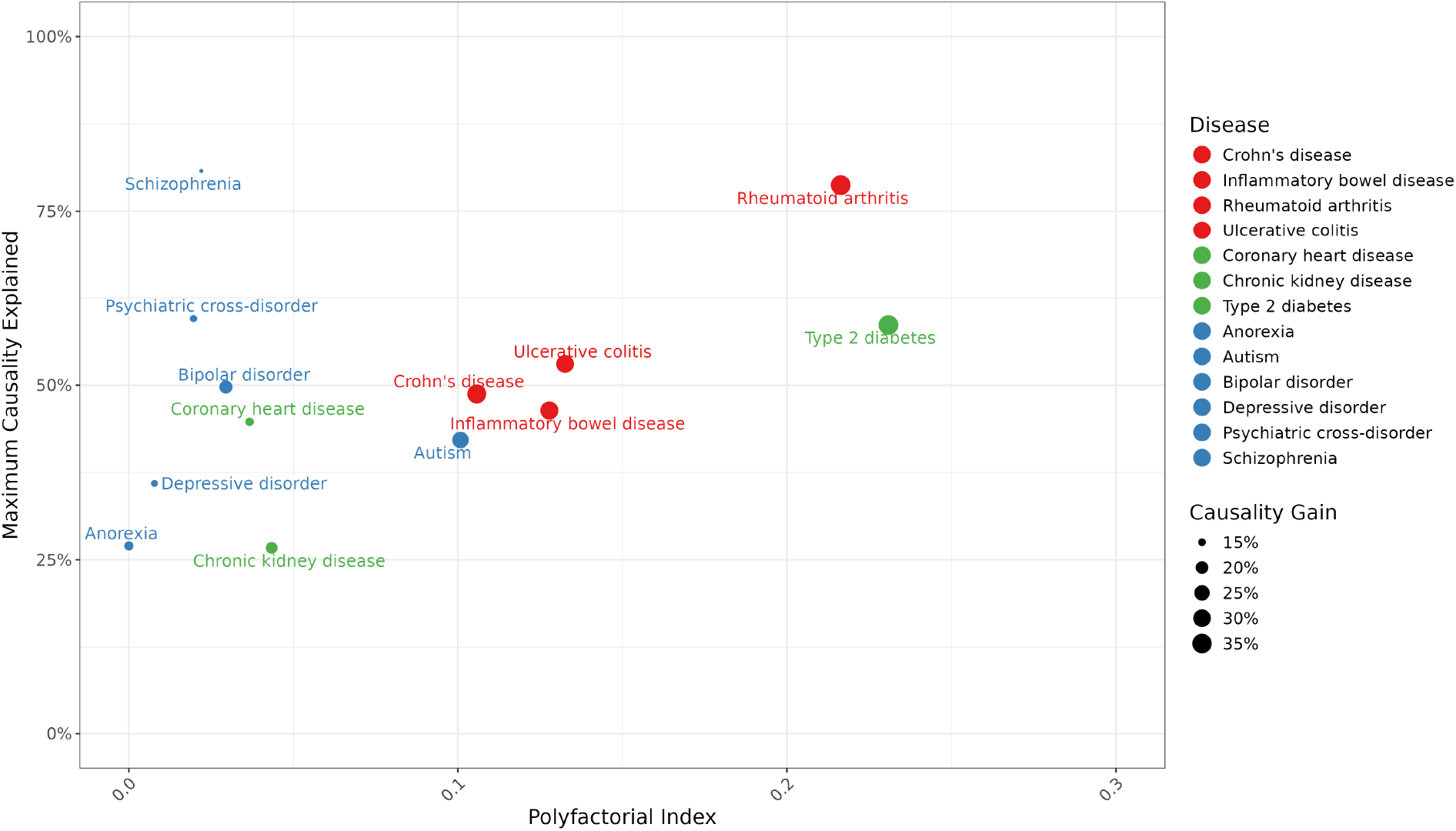
Representation of the maximum causality explained (y-axis) according to the polyfactorial index (x-axis) for 13 diseases in 3 phenotypic categories represented by the colors. The size of the dot is proportional to the causality gain.

**Figure 4.**
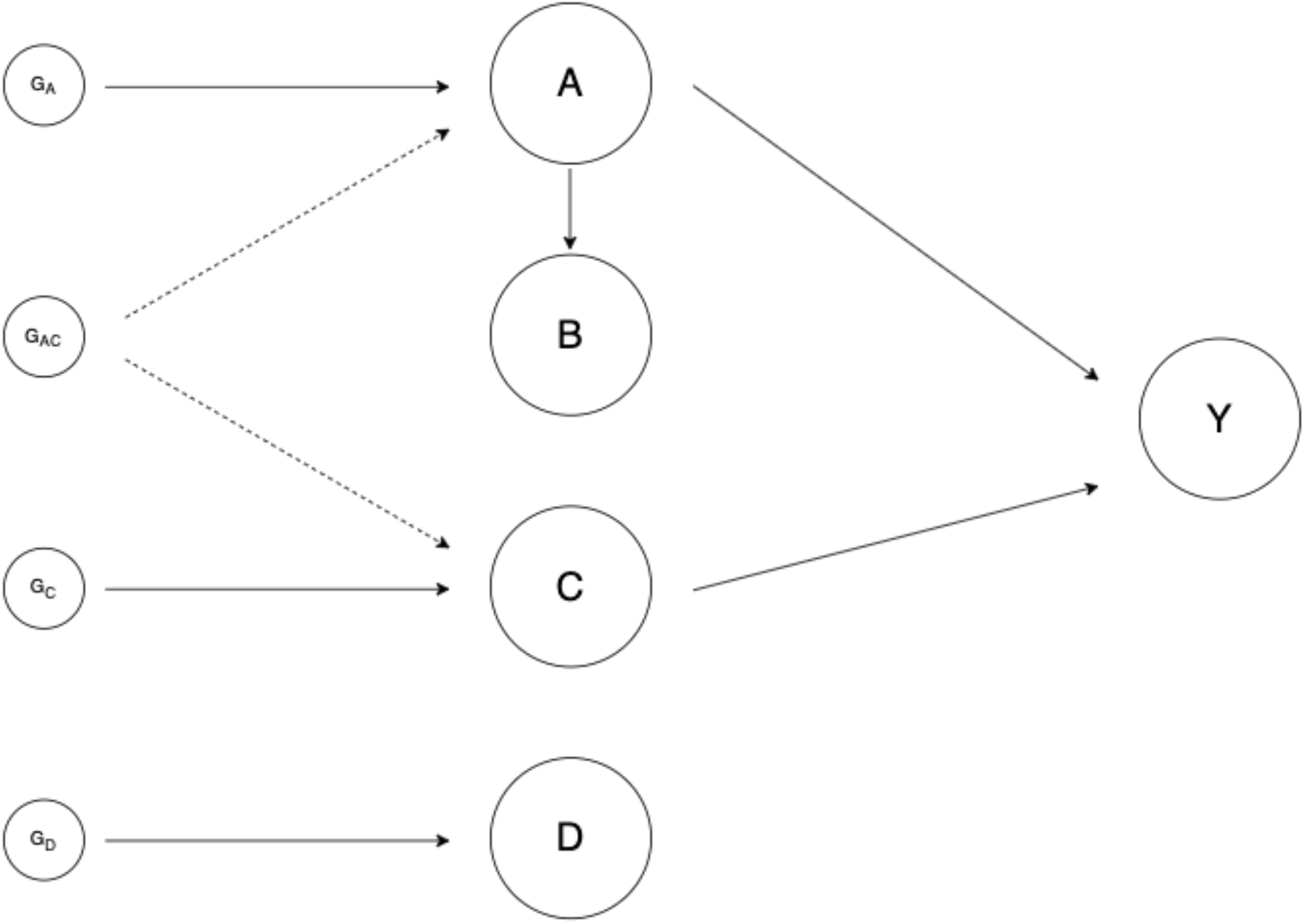
Simulation framework. Y is the outcome trait. A, B, C, and D represent sets of exposures with causal relationships between the exposure sets and the outcome represented by arrows. G_A_, G_B_, G_C_, and G_D_ represent the set of genetic variants that are used as instrumental variables for exposures A, B, C, and D respectively. If horizontal pleiotropy is included in the simulation scenario, a given proportion of instrumental variables have significant effects both on exposures from sets A and C.

To complement the interpretation of the results in terms of causal architecture for our 13 complex diseases, we investigated the link between our causality explained model and the heritability estimated from twin studies that we could retrieve from the literature. Interestingly we could observe a tendency for more heritable traits to reach a higher maximum causality explained. Finally, schizophrenia was a particularly interesting disease according to our causality explained model because of a combined very high maximum causality explained as well as a very low causality gain. The causality gain is the difference between the causality explained by the first Phecode and the maximum causality explained, which meant that the first PheCode already explained a large proportion of causality for schizophrenia (66.4%). Looking more closely at the PheCodes, which are ordered by their individual level of causality explained in the model, the first PheCodes as well as the following few ones were all closely related to celiac disease. The relation between celiac disease and schizophrenia has been established^22^ and gluten-free diets can be recommended for schizophrenic patients^23^. It is worth mentioning that as in simulations, the trajectories and metrics, using the alternative method to select relevant principal components, were highly similar (Supplementary Figures 10 and 11).

Finally, we were also interested in comparing the causality explained trajectories with traits that were expected to have very different causal architectures than the 13 complex diseases. First, we looked at the causality of 6 brain volume traits from the ENIGMA consortium, specifically the mean volume of the accumbens, caudate, hippocampus, pallidum, putamen, and thalamus. As expected, all the trajectories were highly similar with lower maximum causality explained and lower gain compared to all 13 complex diseases (Supplementary Figure 12). Second, we focused on a trait for which the causal risk factor is well identified which is gout caused by an accumulation of uric acid. The causality explained trajectory was not very interpretable for Gout confirming the monofactorial nature of the trait (Supplementary Figure 13), and further validating the interest of our model in studying polyfactorial traits. Additional details are provided on the interpretation of the results for Gout in the Supplementary Results section.

## Discussion

We have introduced the concept of causality explained for complex diseases as well as a causality explained model. This enabled the inclusion of an exhaustive set of potentially causal exposures in a generalization of the MVMR model based on principal component linear regression to account for colinearities in the exposure effect sizes. In simulations, we showed that our model reached a high level of causality explained approaching 100% when all exposures and most IVs are included in the model. The level of causality explained was sensitive to the detection of IVs which is influenced by multiple factors. First, the sample size of the GWAS on the exposures is directly linked to the power to detect IVs, therefore large studies should be used to maximize the causality explained. Second, the causality explained is strongly influenced by pleiotropy, whether it is horizontal pleiotropy when genetic variants are causal to multiple exposures or vertical pleiotropy when exposures, including non-causal exposures, are causal to each other. Since pleiotropy is widespread^3,16,17^, this is a major innovation of our causality explained model.

On real data, we chose a highly comprehensive set of exposures from a PheWAS on UK Biobank PheCodes which provided us with a well-powered set of about 222 exposures and studied the causality explained of 13 complex diseases. First, we focused on the causality explained for CAD. Our results revealed that an exhaustive set of exposures explained more than twice the amount of causality explained by the known causal risk factors jointly. It is worth mentioning that for CAD, some individual risk factors such as cholesterol-related PheCodes still explained individually a comparatively large proportion of causality around 16% which is in line with both the preventive and therapeutic development around cholesterol and lipids in CAD.

Here, to explain these results, we propose the “omnicausal model” and further distinguish between core and peripheral causal risk factors that explain respectively larger and smaller portions of causality similarly to the omnigenic model^4^. We observed that known causal risk factors explain a larger portion of causality individually, and we hypothesize that these traits represent core causal risk factors. When adding the PheWAS traits, the causality explained reached almost 45%, which we hypothesize are peripheral traits that contribute a small amount to the causal model, but collectively explain a large portion.

In addition, applying the causality explained model to 13 complex diseases into three broad categories (*i*.*e*. metabolic, inflammatory, psychiatric diseases), with the exhaustive set of PheCodes as exposures, led to revealing different causal architectures and corroborated our hypothesis of the omnicausal model for complex diseases.

Nevertheless, the model that we have proposed suffers from several limitations. First, we have shown in our simulations, that our model, is strongly influenced by the set of genetic variants that serve as IVs. Although we used the UK Biobank which is a very well-powered resource for our exposures, we are necessarily missing some IVs because of lack of power. In addition, we have used exposures with effect sizes estimated from the same sample, which could also reduce the power to identify IVs. However, these limitations are more related to the exposures than the outcome as we have shown in simulations. Importantly, when applying our model to the set of 13 complex diseases and using the same exhaustive set of exposures (except for disease-redundant exposures), all 13 models presumably suffer from the same limitations, and hence we still expect different levels of causality explained to be comparable. In addition, it is important to mention that all the results of our causality explained model, as well as the metrics used to interpret the trajectories, are highly dependent on the set of exposures. We do hope to come to robust conclusions by using a phenome-wide set of exposures but at the same time we might have observed inflated causality explained for complex diseases because we used PheCodes which are medical phenotypes, and although we excluded Phecodes redundant with the disease outcome, PheCodes might be more prone to recapitulate complex diseases leading to higher causality explained. In addition, our causality explained model does not allow to pinpoint individual risk factors and we have to use a strategy to account for colinearities in the exposure effect sizes. We chose to use principal component analysis, however selecting the relevant number of principal components is a well-known issue and by using two distinct strategies we do hope to mitigate the risk of biases. It is worth noting that this strategy seemed to be ok for polyfactorial traits however as we have shown it is limited for monofactorial traits (see Supplementary results for details). Another limitation is that currently, we don’t have an understanding of the relationship between the heritability and the causality explained of different diseases. We could hypothesize that the causality explained is the part of heritability due to vertical pleiotropy, *i*.*e*. the genetic effects on disease that are mediated through the risk factors. Future work is warranted to better understand this relationship. Furthermore, we included phenome-wide exposures that span a variety of risk factors which can be confounded with unmeasured factors. This can result in omitted-variable bias and could result in measuring the causality explained by both included and confounded risk factors. We believe that this is not an issue in our case since we do not seek to pinpoint individual causal risk factors but instead intend to have the most comprehensive set of exposures possible. Importantly, we show in our simulations that this omitted-variable bias does not inflate the causality explained statistic. Finally, there could be a lot of heterogeneity in genetic effects across populations that were not taken into account in this study because we mostly focused on European samples.

In conclusion, we have proposed a new concept of causality explained through the causality explained model and associated metrics to quantify the causality of a set of risk factors on disease. Importantly, this work proposes a new model called the “omnicausal model” with core and peripheral causal risk factors. We have shown that at least applied to CAD, a large proportion of missing causality is explained by many non-traditional traits with individually small effects on disease that collectively have a large effect on the causal variance of disease. Furthermore, when applied to several complex diseases, a large number of risk factors collectively explain a large portion of the causal variance to these diseases.

## Methods

### Multivariable Mendelian randomization

Multivariable Mendelian Randomization (MVMR) is a method that generalizes the classical single-variable MR (SVMR) framework, where the causal effect for one exposure on one outcome is modeled, by jointly estimating the causal effect of several exposures on disease outcome^14,15^. MVMR is therefore performed in a multivariate linear regression context (with an intercept fixed to zero) where the response variable is the estimated effect sizes on the outcome and the predictors are the estimated effect sizes on the included exposures. Importantly, all the genetic variants associated with at least one exposure, denoted instrumental variables (IVs) are included in the MR model. MVMR can be implemented using summary statistics estimates of the association between the genetic variant *j* and the outcome *ω*_*j*_, and multiple exposures denoted 1 to 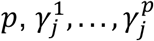 by fitting the following model:

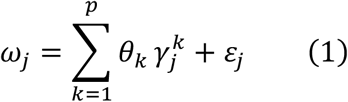

This is a direct generalization of the Inverse Variance Weighted (IVW) estimation framework since the multivariate regression included weights depending on the standard error of the variant effect sizes on the outcome.

### Concept of causality explained

We define the causality explained as the variance explained by one or more risk factors on disease in a MR framework. Then, with *n* the number of IVs and *p* the number of exposures included in the model, we define the causality explained as:

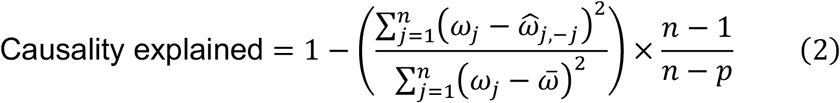

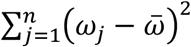 is the total sum of squares and *n* − 1 and *n* − *p* correspond to the total and residual degrees of freedom respectively. 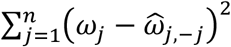 refers to the predicted error sum of squares which is a form of cross-validated predicted error in a leave-one-out procedure where each IV is sequentially omitted to compute the prediction error^24^. This metric would correspond to a predicted and adjusted *R*^2^.

For a given set of exposures, we define a panel of IVs as the set of genetic variants associated with at least one exposure in the panel. It is possible to include genetic instruments associated with more than one exposure, given that all those exposures are included in the model, which we address by using an exhaustive panel of exposures.

Notably, to conduct an MVMR analysis, it is necessary to have at least as many genetic instruments as there are exposures. A proof-of-concept paper has shown that it is possible to combine several exposures into a MVMR model despite colinearities between exposures^25^, in the paper, principal component analysis (PCA) has been successfully applied to the summary statistics of 14 anthropometric traits to study their causal effect on obesity. In addition, a sparse PCA version has applied to 97 highly correlated lipid metabolites in a MVMR setting^26^. To address the concern of potential overfitting we also conduct principal component analysis on the matrix of genetic instruments. We use the resulting PC loadings in equation (1). Selecting the correct number of PCs is a well-known issue in PCA, therefore, we consider two distinct PC selection method: using all PCs, selecting PCs with eigenvalues 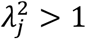 and based on 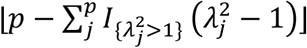 ^27^

### Causality explained model and associated metrics

The procedure to fit the causality explained model involves three key steps. 1) In the preliminary steps, relevant instrumental variables (IVs) for each exposure are selected, ensuring a set of independent IVs across all exposures. Individual levels of causality explained are then calculated for each exposure which are then ordered them according to their decreasing levels of individual causality explained. 2) The causality explained model is initialized using the causality explained by the first exposure. Exposures are then sequentially added to the model, and principal component (PC) analysis is performed to account for collinearities among their effect sizes. The appropriate number of PCs is selected, and the model is fit to compute the causality explained. 3) We model the causality explained trajectory. In addition to the causality explained trajectory, we compute three metrics to characterize the trajectory: the maximum causality explained, the causality gain as the difference between the causality explained by the first exposure and the maximum causality explained, and the polyfactorial index. The polyfactorial index quantifies the maximum gain in causality explained by comparing the observed trajectory of causality explained with its theoretical trajectory. The theoretical trajectory assumes a steady increase from the percentage of causality explained by the first exposure up to 100%. In theory, the polyfactorial index could approach a value of 1, indicating that the causality explained rises sharply with the addition of only a few exposures. Conversely, the minimum value of the index would be close to zero or even negative, signifying that the causality explained increases gradually, with each additional exposure making a relatively equal contribution to reaching the maximum causality explained.

### Simulation framework

We define four different types of exposures.

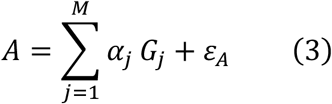

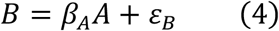

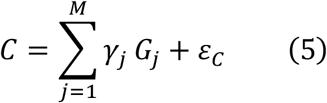

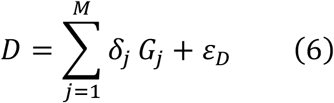

The outcome of interest *Y* is defined as:

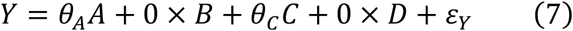

We generate genotypes *G*_*j*_ by first simulating 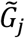 from a binomial distribution *B*(2,0.3) and then standardizing by 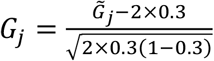 to make them have mean 0 and variance 1, which means they have a minor allele frequency of 0.3. We generate data from equations (3), (4), (5), (6) and (7) using 50,000 independent genetic variants. We fix the total number of exposures to 100, and we vary the number of exposures that belong to type *A, B, C* and *D*. In the case of no horizontal pleiotropy, we simulate each exposure from 100 genetic variants that are independent across exposures. In the case of horizontal pleiotropy, we vary the proportion of shared IVs across exposures in sets *A* and *C*. For each exposure in set *C*, a fraction of its causal genetic variants overlaps with the causal genetic variants from exposures in set *A*, while keeping mutually exclusive exposures in *C*. We vary the proportion of shared genetic variants: 0%, 20%, 50% and 80%. The error terms *ε*_*A*_, *ε*_*B*_, *ε*_*C*_, *ε*_*D*_ and *ε*_*Y*_ are independent and normally distributed with mean 0. For simplicity, we chose the variance error terms so that *A, B, C, D* and *Y* have all have variance 1. And *α*_*j*_, *β*_*j*_, *γ*_*j*_ and *δ*_*j*_ are the direct genetic effect of variant *j* on *A, B, C*, and *D*, respectively. These effects are independent and normally distributed with mean 0 and variances set to either 0.008 or 0.004. This choice ensures comparable power to detect IVs across different simulation scenarios, as we do not compare scenarios that use different variances. *B* is in vertical pleiotropy with *A*, and *β*_*A*_ is the causal effect of *A* on *B*, however *B* is not causal of *Y*. When we have more than one exposure in *A*, for each exposure in *B* we sample one exposure in *A* to be in vertical pleiotropy. *θ*_*A*_ and *θ*_*C*_ are nonzero causal effects of the exposures *A* and *C* on *Y*. That is the set of exposure in *A* and *C* are always causal to *Y*, the set of exposures in *B* and *D* are not causal to *Y*, but *B* is in vertical pleiotropy with *A* and *D* is independent on the others. To induce correlated pleiotropy, we simulated a confounding variable *U* that links each exposure with *Y*. For every genetic variant that exerts a direct effect on any exposure, an independent genetic effect on *U* was also assigned. These genetic effects on *U* were independently drawn from the same normal distribution as the direct effects on the exposures. Subsequently, the confounder *U* was modeled to contribute 1% of its effect to the respective exposure and 0.*5*% to the outcome.

### Application to Coronary Artery Disease and complex diseases

We applied the causality-explained model to coronary artery disease (CAD) as the outcome in MR testing. The GWAS summary statistics for coronary artery disease (CAD) were retrieved from the CARDIoGRAMplusC4D Consortium^21^. As exposures in MR testing, GWAS summary statistics were retrieved from a phenome-wide association study (PheWAS) on 408,961 individuals from the UK Biobank on PheCodes derived from electronical medical records. The summary statistics of the PheWAS were obtained using SAIGE that control for the inflation of type 1 error due to extreme imbalance of cases and controls^28^. Five PheCodes that are redundant of CAD were excluded (Supplementary Table 2). We retained 222 PheCodes with at least one instrumental variable, the total of IVs was then 5,314.

Furthermore, we applied the causality-explained model to a range of complex diseases for which summary statistics GWAS data were retrieved from the literature. For each of these diseases, redundant phecodes were excluded (Supplementary Table 2). The included diseases comprise Alzheimer’s disease (AD)^29^, anorexia nervosa (AN)^30^, autism spectrum disorder (ASD)^31^, bipolar disorder (BIP)^32^, Crohn’s disease (CD)^33^, chronic kidney disease (CKD)^34^, inflammatory bowel disease (IBD)^33^, major depressive disorder (MDD)^35^, psychiatric cross disorder (PGC_cross)^31^, rheumatoid arthritis (RA)^36^, schizophrenia (SCZ)^37^, type 2 diabetes (T2D)^38^ and ulcerative colitis (UC)^33^. In addition, the brain volume traits were retrieved from the ENIGMA consortium^39^ and the gout summary statistics were retrieved from the Global Urate Genetics Consortium^40^.

All GWAS summary statistics were preprocessed to ensure comparability of genome built, strand, reference and alternate allele.

## Data Availability

The summary statistics of the PheWAS (obtained using SAIGE) and the GWAS summary statistics for coronary artery disease (retrieved from the CARDIoGRAMplusC4D Consortium) are available on Zenodo. Scripts for reproducibility are available on github. Other summary statistics are openly available online as sourced in the manuscript.

https://zenodo.org/records/14860110

https://github.com/martintnr/CausalityExplained

## Data and code availability

The code to run the causality explained model is implemented in R and is available on github, coupled with examples of data to run the causality explained model on Zenodo.

## Authors contributions

CM-L, contributed to study conception, data analysis, interpretation of the results and drafting of the manuscript; MT contributed to study conception, data analysis, interpretation of the results, drafting and revision of the manuscript; GR contributed to study conception, interpretation of the results and revision of the manuscript; and RD and MV contributed to study conception, interpretation of the results and critical revision of the manuscript.

