## Supplementary Material for "The omnicausal model reveals the highly polyfactorial nature of complex diseases"


#### Supplementary Figures

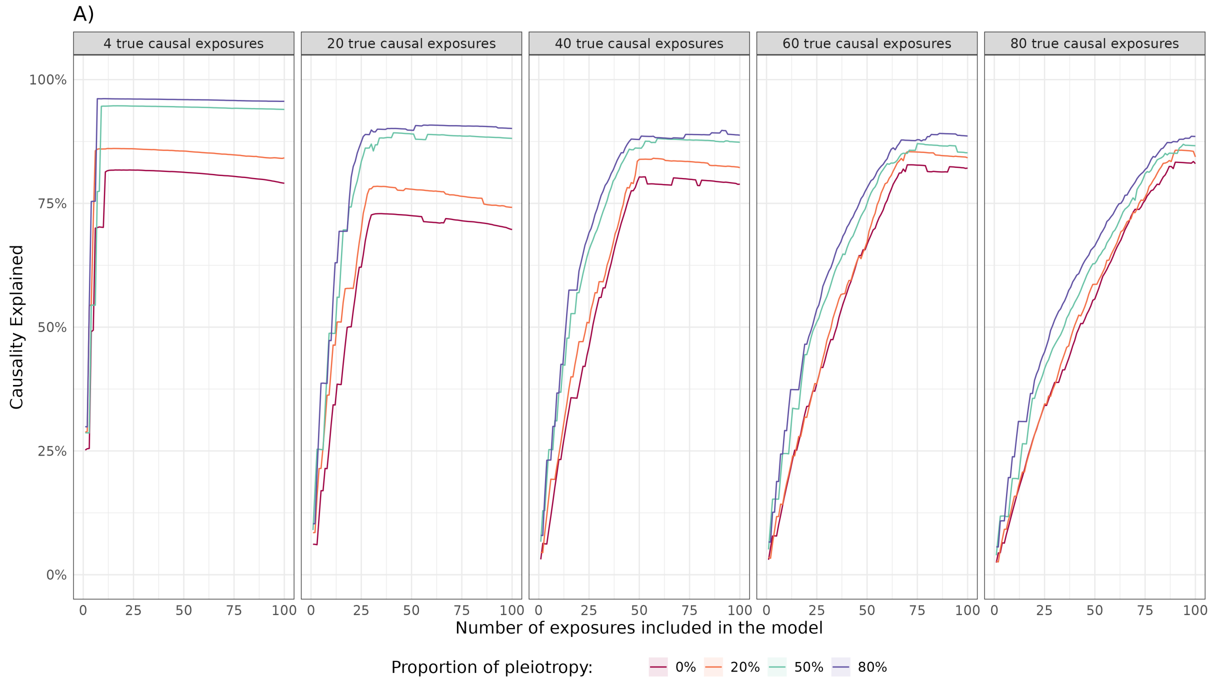


***Supplementary Figure 1.*** *Behaviour of the causality explained model in one specific simulation. The y-axis represents the evolution of the causality explained according to the size of the multivariable Mendelian randomization model (x-axis), by sequentially including exposures into the model. At each step principal component analysis is used to account for colinearities. Horizontal and vertical panes respectively distinguish the true number of causal exposures, and two levels of per-variant heritability. Colors represent different pleiotropic architectures induced by the proportion of causal variants with pleiotropic effect on at least two exposures.*

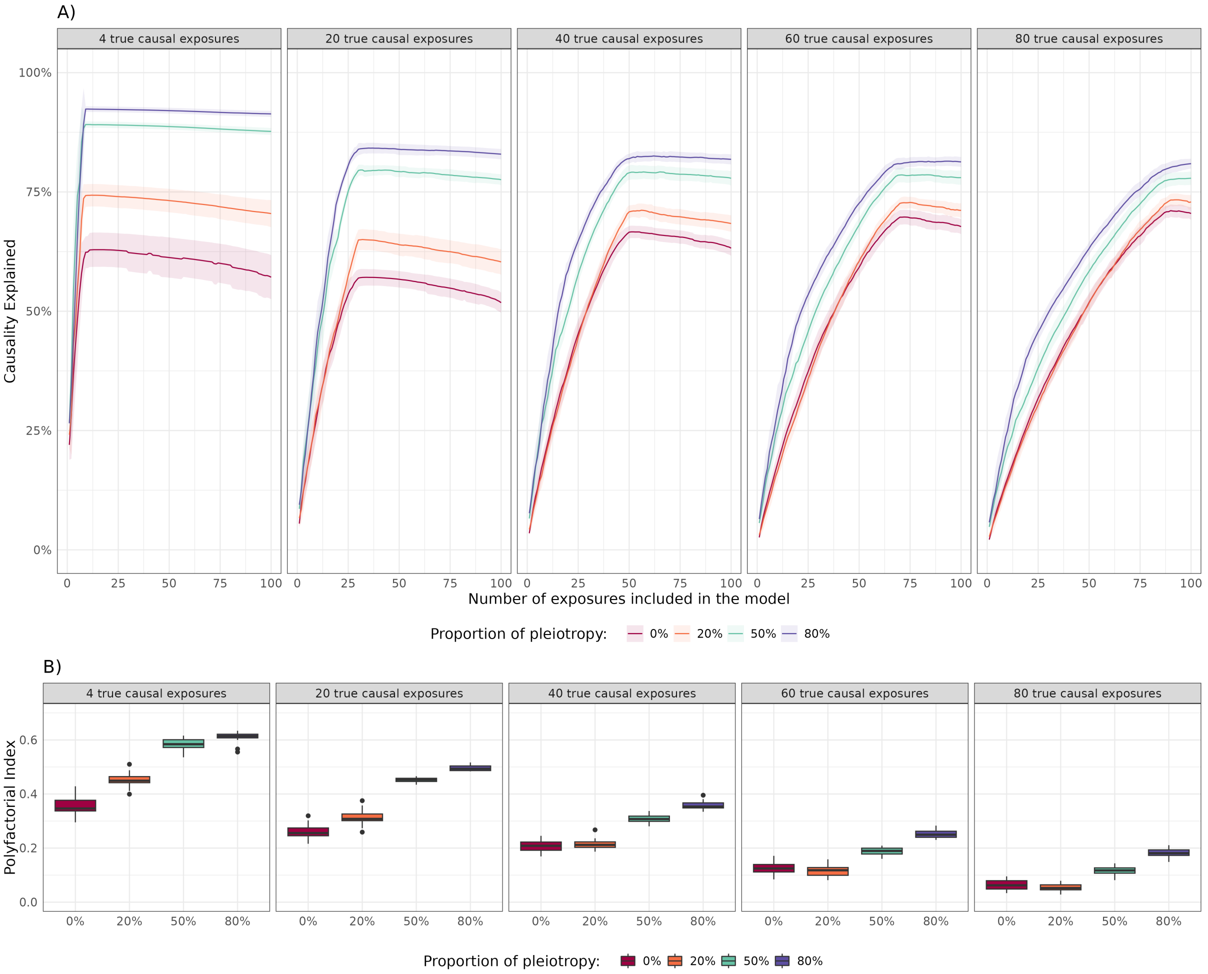


***Supplementary Figure 2.*** *Causality explained and polyfactorial index in case of low per-variant heritability. In panel* ***A)****, the y-axis represents the evolution of the average causality explained and standard error over 20 replicates, according to the size of the multivariable Mendelian randomization model (x-axis), by sequentially including exposures into the model. At each step principal component analysis is used to account for colinearities. In panel* ***B)****, the y-axis represents the polyfactorial index that characterizes the trajectory of causality explained when adding exposures one by one according to the causal variants pleiotropy (x-axis). In both panels, the panes distinguish the true number of causal exposures, and colors represent different pleiotropic architectures induced by the proportion of causal variants with pleiotropic effect on at least two exposures.*

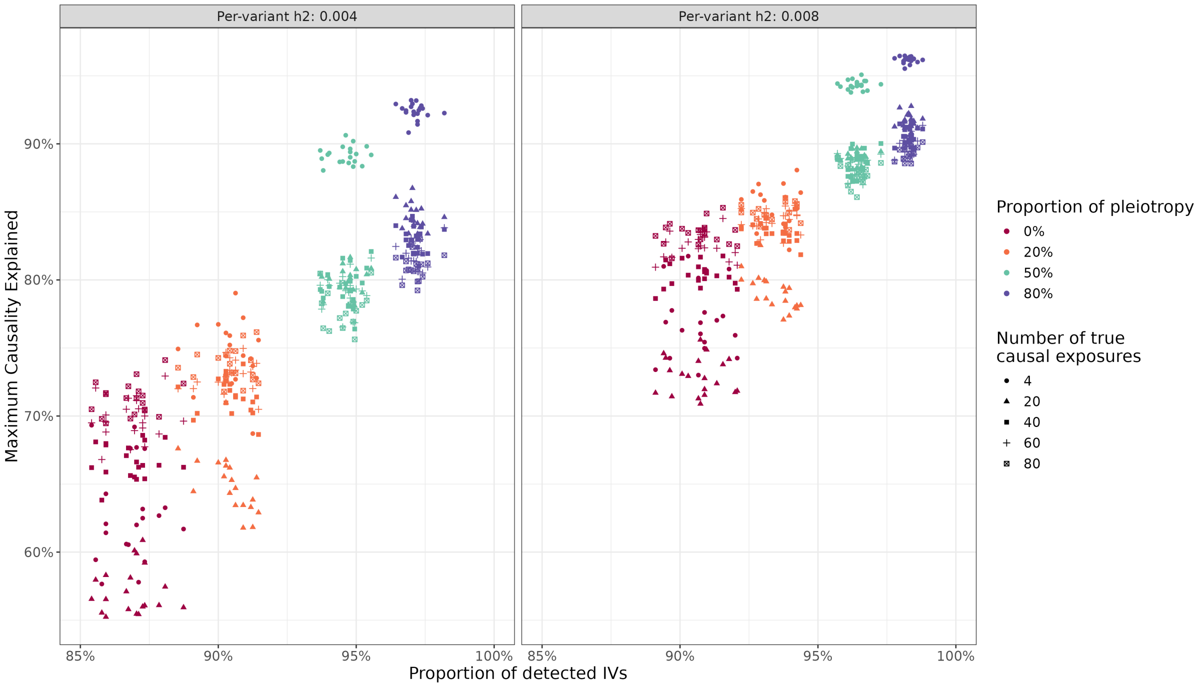


***Supplementary Figure 3.*** *Evolution of the top causality explained on the y-axis, according to the proportion of detected instrumental variables per exposure on the x-axis. The panes distinguish two levels of per-variant heritability. Colors represent different pleiotropic architectures induced by the proportion of causal variants with pleiotropic effect on at least two exposures. Shapes represent the number of true causal exposures. Results for 20 replicates for each scenario are represented.*

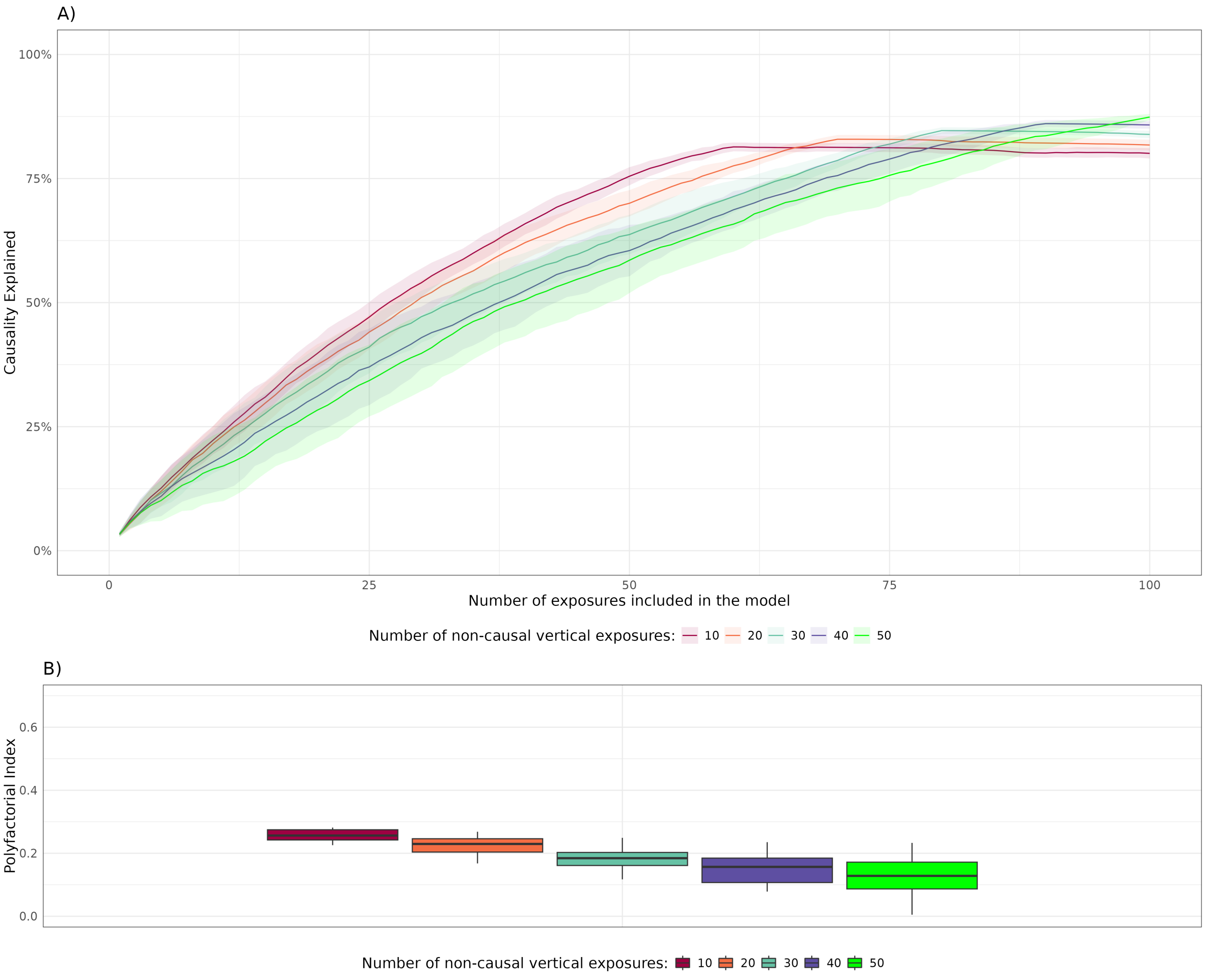


***Supplementary Figure 4.*** *Assessment of the effect of vertical pleiotropy of non-causal exposures on causality explained.* ***A)****, the y-axis represents the evolution of the average causality explained and standard error over 20*

replicates, according to the size of the multivariable Mendelian randomization model (x-axis), by sequentially including exposures into the model. At each step, principal component analysis is used to account for colinearities. ***B)***, *the y-axis represents the polyfactorial index that characterizes the trajectory of causality explained when adding exposures one by one according to the causal variants pleiotropy (x-axis). The colors represent the number of non-causal exposures in vertical pleiotropy. 50 exposures are causal, and 10 and 40 belong to group $A$ and group $C$ respectively. 50 exposures are non-causal and belong to either group $B$ which is in vertical pleiotropy with group $A$, or group $D$ which is not in vertical pleiotropy. The number of non-vertical exposures in vertical pleiotropy is therefore the number of exposures in group $B$.*

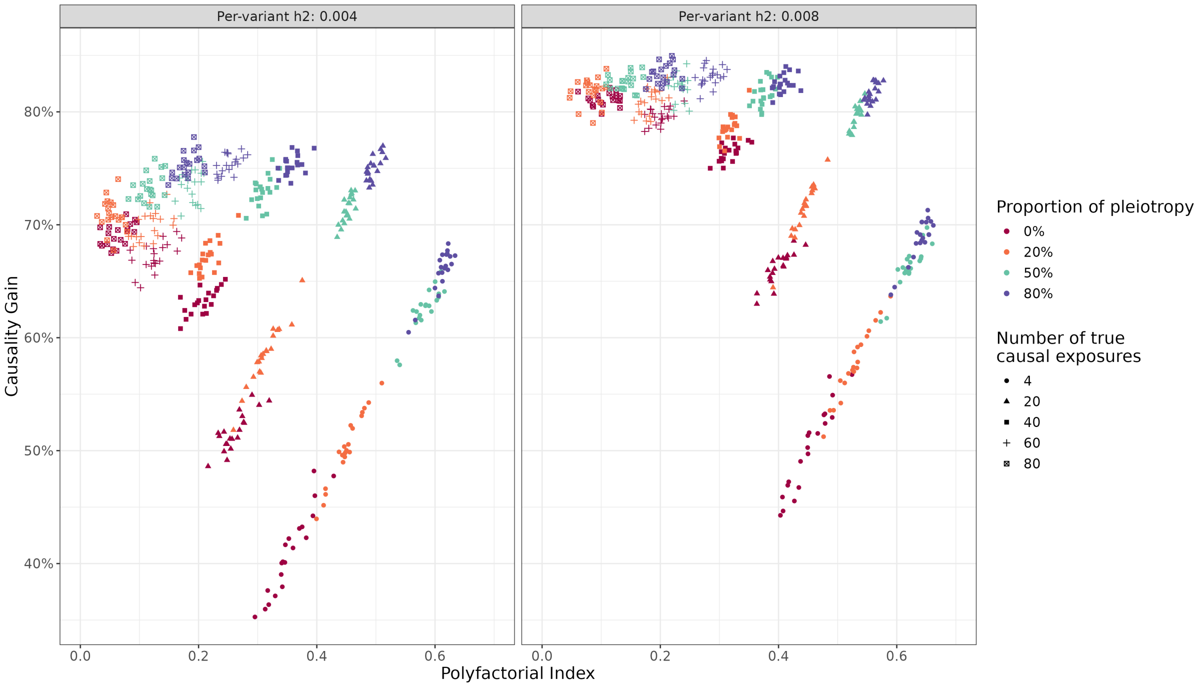


***Supplementary Figure 5.*** *Evolution of the causality gain on the y-axis, according to the polyfactorial index on the x-axis. The panes distinguish two levels of per-variant heritability. Shapes represent the true number of causal exposures. Colors represent different pleiotropic architectures induced by the proportion of causal variants with pleiotropic effect on at least two exposures. Results for 20 replicates for each scenario are represented.*

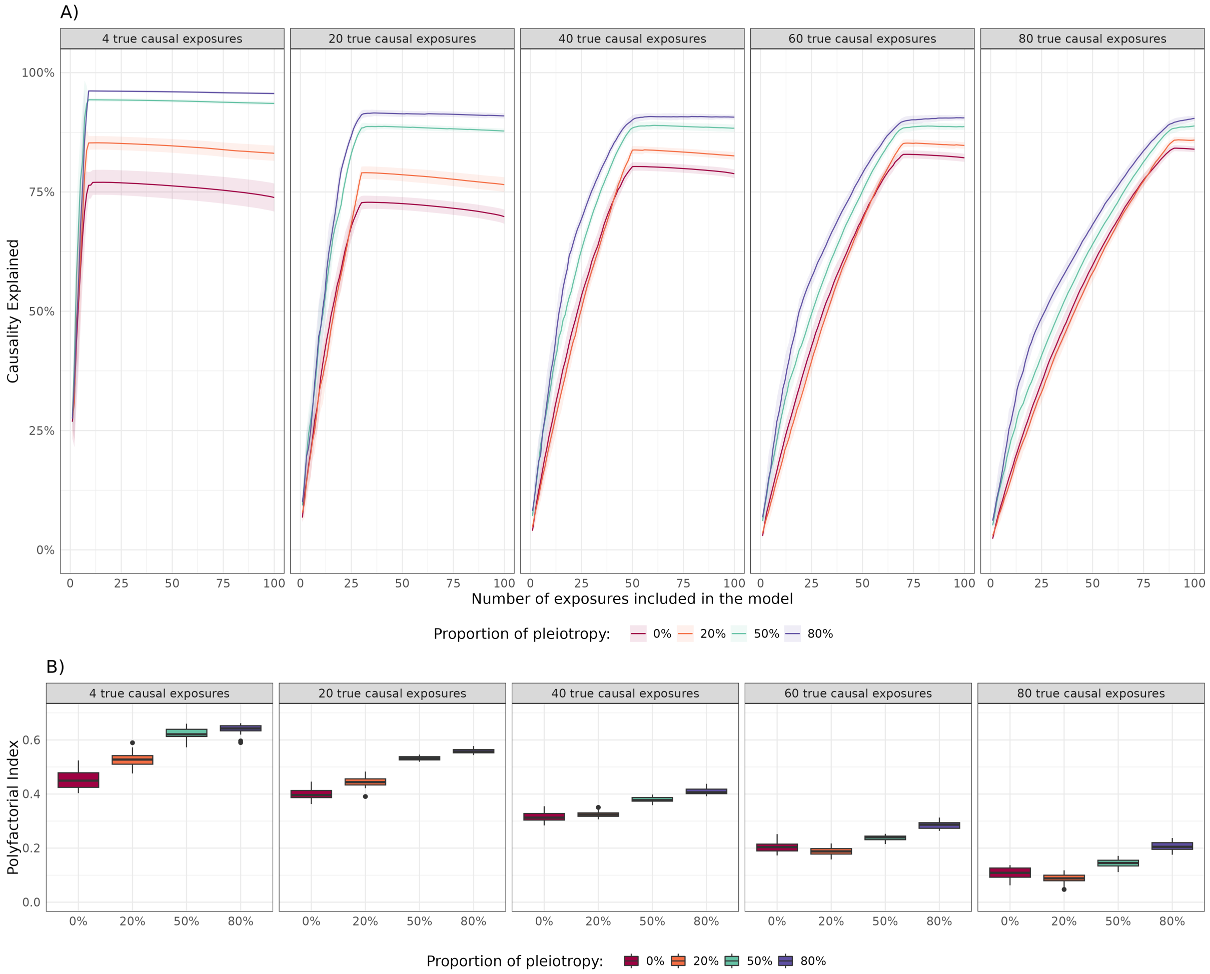


***Supplementary Figure 6.*** *Evaluation of the omnicausal model in simulation with an alternative principal component selection procedure. In panel* ***A)****, the y-axis represents the evolution of the average causality explained and standard error over 20 replicates, according to the size of the multivariable Mendelian randomization model (x-axis), by sequentially including exposures into the model. At each step principal component analysis (PCA) is used to account for colinearities. Here, PCs with eigenvalues greater then 1 in the standardized PCA were considered (See Methods for details). In panel* ***B)****, the y-axis represents the polyfactorial index that characterizes the trajectory of causality explained when adding exposures one by one according to the causal variants pleiotropy (x-axis). In both panels, the panes distinguish the true number of causal exposures, and colors represent different pleiotropic architectures induced by the proportion of causal variants with pleiotropic effect on at least two exposures.*

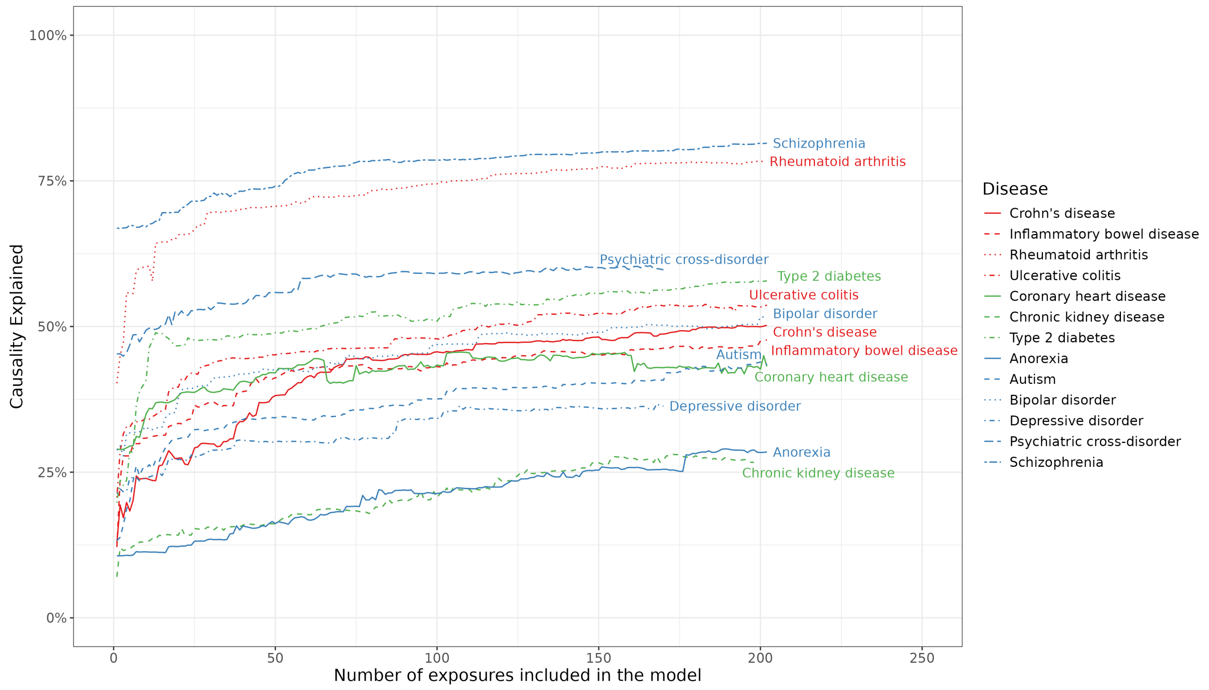


***Supplementary Figure 7.*** *Evolution of the causality explained (y-axis) in 13 complex diseases according to the size of the multivariable Mendelian randomization model (x-axis) by sequentially including PheCodes from the UK Biobank, ordered by individual causality explained, into the model. The PheCodes used in this analysis represent the subset of common PheCodes for all 13 diseases, i.e after filtering out PheCodes that are redundant with any of the diseases. Diseases are color-coded according to broad categories.*

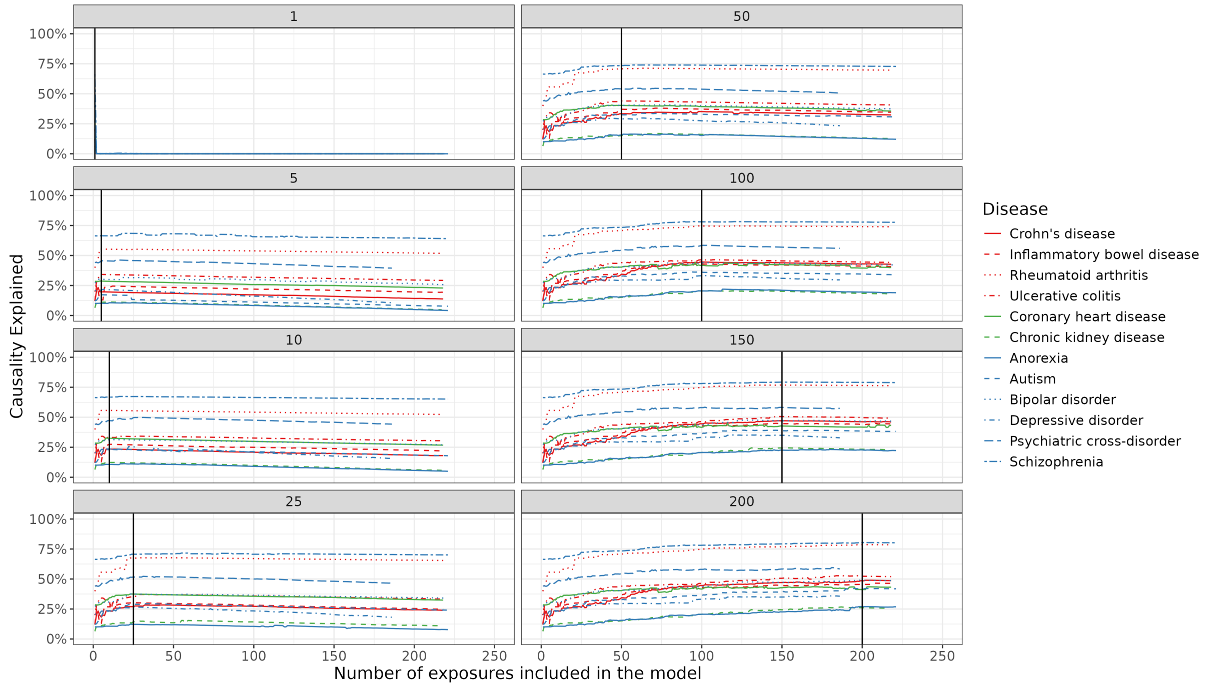


***Supplementary Figure 8.*** *Evaluation of the omnicausal model in 13 complex diseases with permuted PheCode summary statistics from the UK Biobank. In each panel, the trajectory of the causality explained (y-axis) is represented according to the size of the multivariable Mendelian randomization model (x-axis), by sequentially including PheCodes into the model. The vertical black line indicates the PheCode from which the effect sizes are permuted for subsequent PheCodes included in the model.*

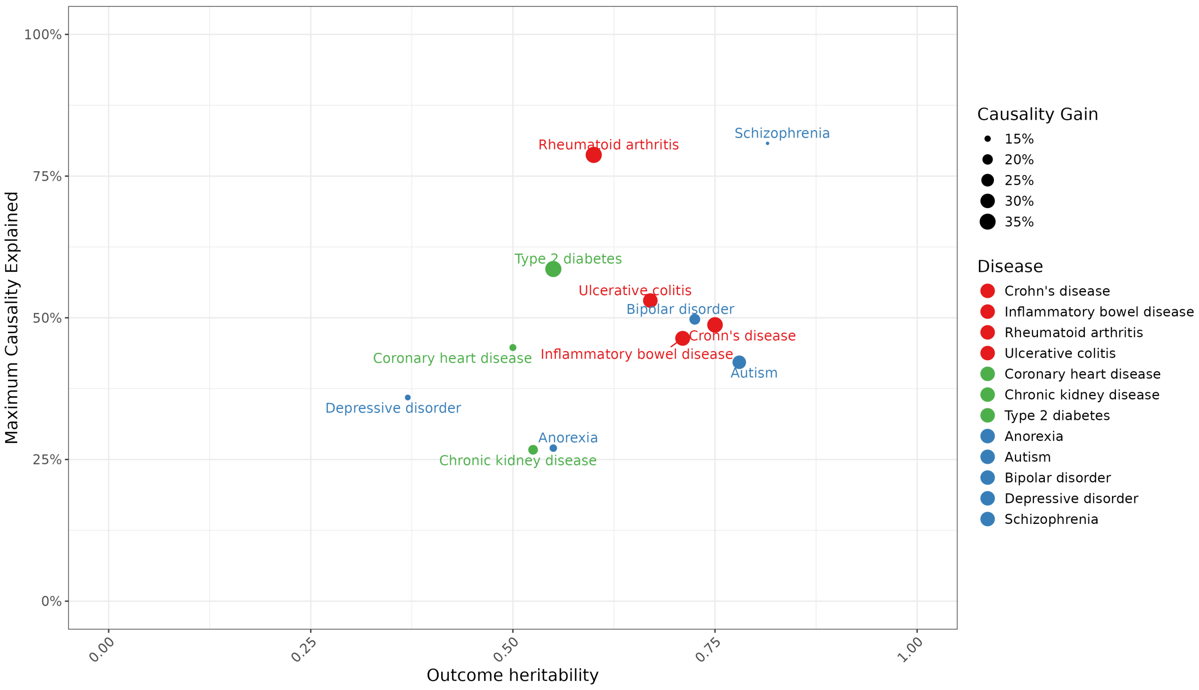


***Supplementary Figure 9.*** *Evaluation of the omnicausal model characteristics in 13 complex diseases using PheCodes from the UK Biobank compared to disease heritability. The maximum causality explained (y-axis) is represented according to average heritability estimates from the literature (x-axis). The size of the dots is proportional to the causality gain that represents the difference between the causality explained by the first PheCode and the maximum causality explained.*

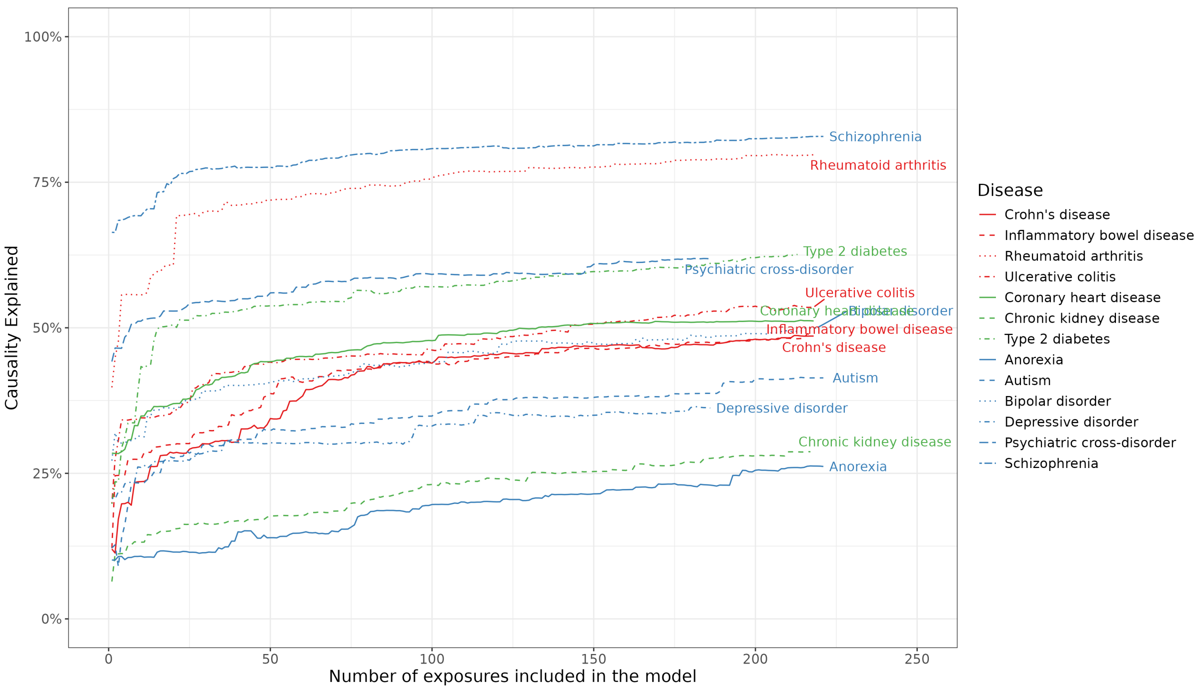


***Supplementary Figure 10.*** *Evaluation of an alternative principal component selection method in the causality explained model. The causality explained (y-axis) in 13 complex diseases is represented according to the size of the multivariable Mendelian randomization model (x-axis) by sequentially including PheCodes from the UK Biobank, ordered by increasing individual causality explained, into the model. Diseases are color-coded according to broad categories.*

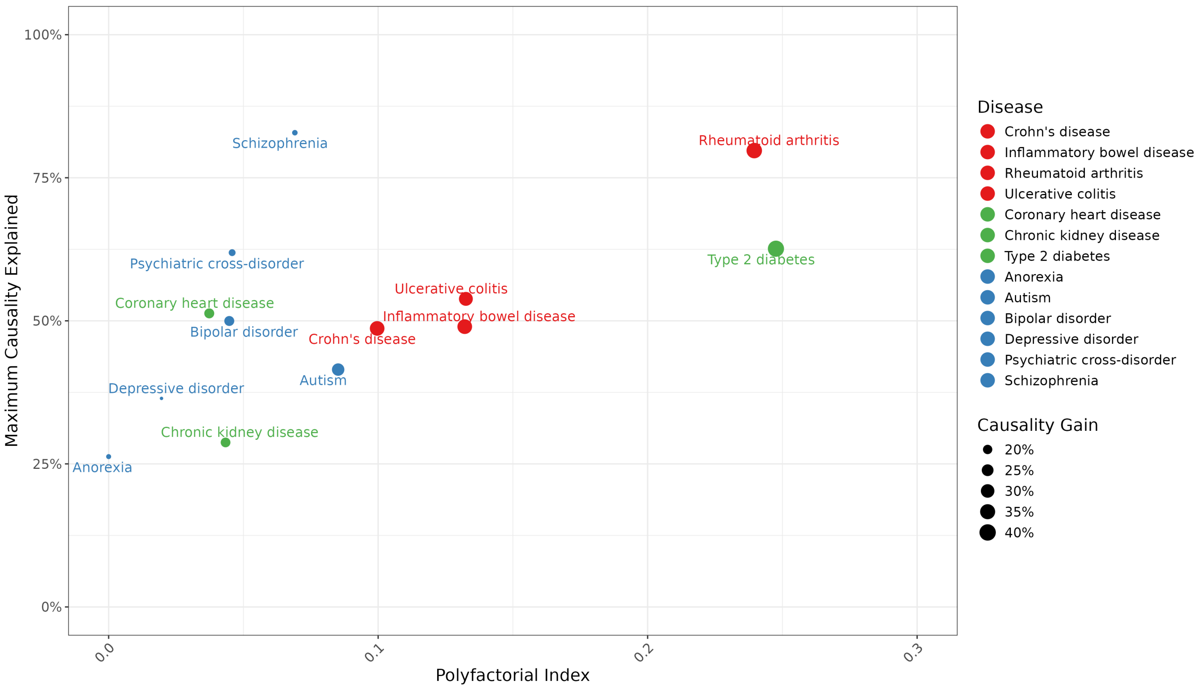


***Supplementary Figure 11.*** *Evaluation of an alternative principal component selection method in the causality explained metrics The maximum causality explained (y-axis) is represented according to the the polyfactorial index (x-axis) for 13 diseases in 3 phenotypic categories represented by the colors. The size of the dot is proportional to the causality gain.*

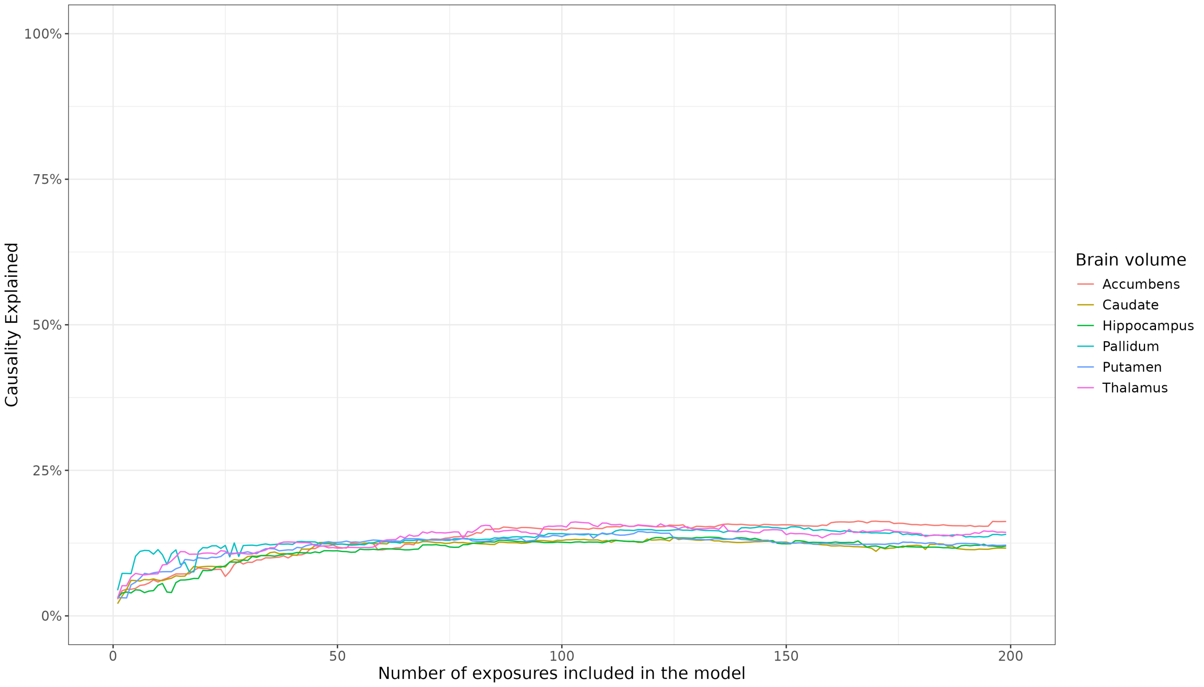


***Supplementary Figure 12.*** *Trajectory of the causality explained (y-axis) according to the size of the multivariable Mendelian randomization model (x-axis), by sequentially including PheCodes into the model for 6 brain traits.*

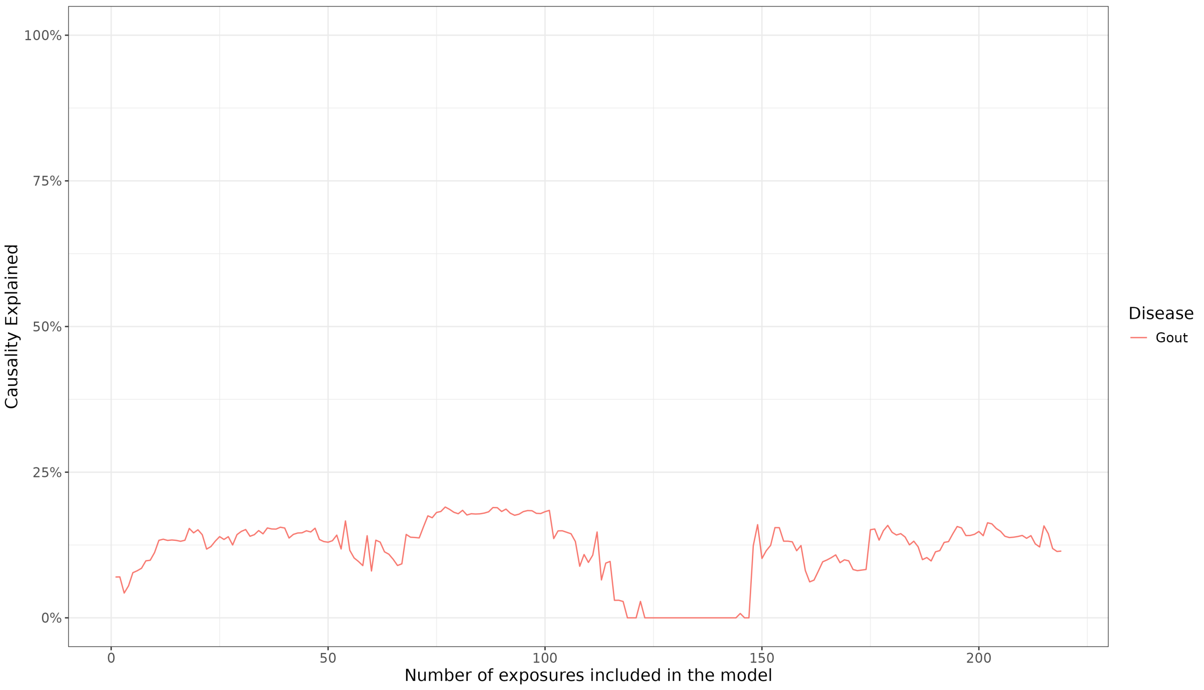


***Supplementary Figure 13.*** *Evolution of the causality explained (y-axis) in for Gout disease according to the size of the multivariable Mendelian randomization model (x-axis) by sequentially including PheCodes from the UK Biobank, ordered by increasing individual causality explained, into the model.*

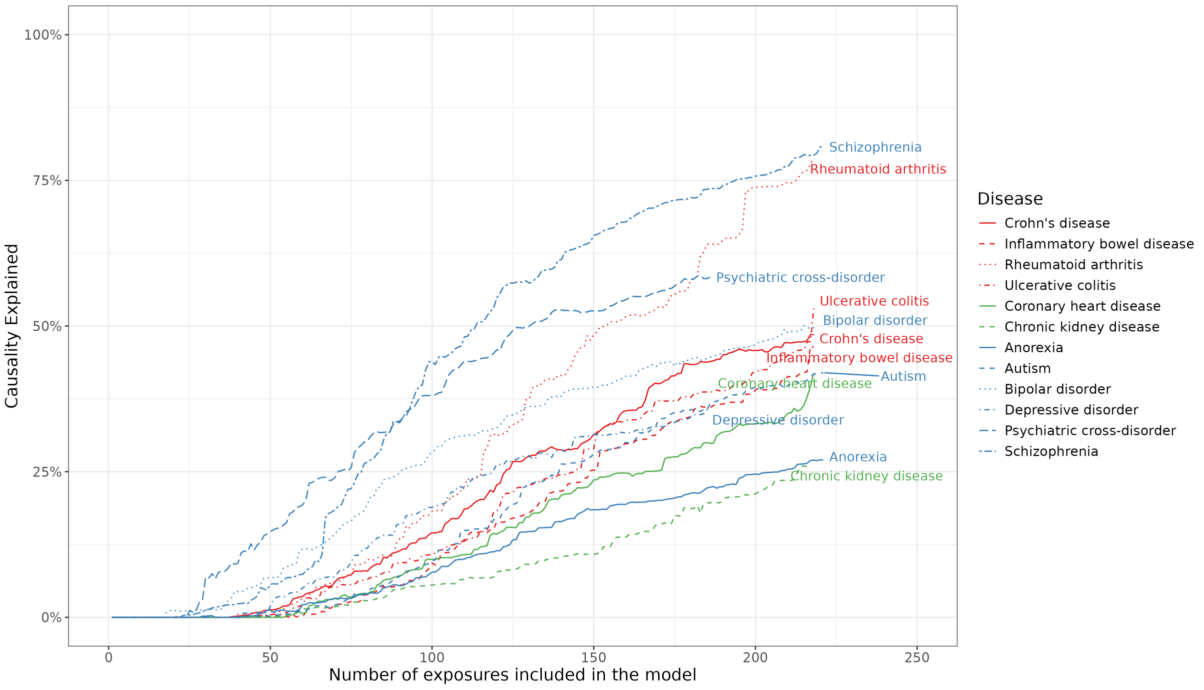


***Supplementary Figure 14.*** *Evolution of the causality explained (y-axis) in 13 complex diseases according to the size of the multivariable Mendelian randomization model (x-axis) by sequentially including PheCodes from the UK Biobank, ordered by decreasing individual causality explained, into the model. Diseases are color-coded according to broad categories.*

### Supplementary Tables

***Supplementary Table 1.*** *Causality explained in coronary artery disease (CAD) in a single-variable Mendelian randomization (SVMR) model for known risk factors individually, or in a multi-variable Mendelian randomization*

*(MVMR) model for all known risk factors or phenome-wide set of PheCodes. CAD summary statistics were retrieved from the CARDIoGRAM consortium.*

| Type of MR model | Included exposures | Causality explained (%) |
|---|---|---|
| SVMR | Hyperlipidemia (272.1) | 16.73 |
| SVMR | Disorders of lipoid metabolism (272) | 16.71 |
| SVMR | Hypercholesterolemia (272.11) | 16.36 |
| SVMR | Hypertension (401) | 10.90 |
| SVMR | Type 2 diabetes (250.2) | 1.70 |
| MVMR | All known clinical risk factors | 17.88 |
| MVMR (Causality explained model) | Phenome-wide PheCodes | 39.34 |

***Supplementary Table 2.*** *List of PheCodes that were excluded from the causality-explained model of the specified complex disease outcome. AD: Alzheimer's disease, AN: anorexia nervosa, ASD: autism spectrum disorder, BIP: bipolar disorder, CD: Crohn's disease, CKD: chronic kidney disease, IBD: inflammatory bowel disease, MDD: major depressive disorder, PGC_cross: psychiatric cross disorder, RA: rheumatoid arthritis, SCZ: schizophrenia, T2D: type 2 diabetes and UC: ulcerative colitis.*

| Disease outcome | PheCode | PheCode Description |
|---|---|---|
| AD | 290.10 | Dementias |
| AD | 290.11 | Alzheimer's disease |
| AN | 305.20 | Eating disorder |
| AN | 305.21 | Anorexia nervosa |
| ASD | 313.30 | Autism |
| BIP | 296.10 | Bipolar |
| CAD | 411.00 | Ischemic Heart Disease |
| CAD | 411.20 | Myocardial infarction |
| CAD | 411.30 | Angina pectoris |
| CAD | 411.40 | Coronary atherosclerosis |
| CAD | 440.00 | Atherosclerosis |
| CD | 555.10 | Regional enteritis |
| CKD | 585.30 | Chronic renal failure [CKD] |
| Gout | 274.10 | Gout |
| Gout | 274.11 | Gouty arthropathy |

| Disease outcome | PheCode | PheCode Description |
|---|---|---|
| Gout | 588.00 | Disorders resulting from impaired renal function |
| IBD | 555.10 | Regional enteritis |
| IBD | 555.20 | Ulcerative colitis |
| IBD | 555.21 | Ulcerative colitis (chronic) |
| MDD | 296.00 | Mood disorders |
| MDD | 296.22 | Major depressive disorder |
| PGC_cross | 295.10 | Schizophrenia |
| PGC_cross | 296.00 | Mood disorders |
| PGC_cross | 296.10 | Bipolar |
| PGC_cross | 296.22 | Major depressive disorder |
| PGC_cross | 313.10 | Attention deficit hyperactivity disorder |
| PGC_cross | 313.30 | Autism |
| RA | 714.10 | Rheumatoid arthritis |
| SCZ | 295.10 | Schizophrenia |
| T2D | 250.20 | Type 2 diabetes |
| UC | 555.20 | Ulcerative colitis |
| UC | 555.21 | Ulcerative colitis (chronic) |

### Supplementary Results

#### The causality explained model is not well-suited for monofactorial traits

As we have commented in the main Results section and as shown in Supplementary Figure 13, the causality explained model is not well-suited for monofactorial traits such as gout which is caused by the accumulation of uric acid. Unsurprisingly, the PheCode that had the highest causality explained is uric acid and as more and more PheCodes were included in the model, we observed large fluctuations in the causality explained. These fluctuations were due to the use of the principal component (PC) step in our methodology. Indeed, relevant PCs are selected for each ensemble of included PheCodes and it could happen that the effect sizes of the main causal risk factor for Gout, *i.e.* uric acid, were not very correlated with the selected PCs resulting in a massive drop in causality explained. Therefore, we recommend using the causality explained model in polyfactorial traits.

#### Importance of the order of the PheCodes in the causality explained model

In our causality explained model, we propose to order the exposure according to their decreasing level of individual causality explained, which allows to model the trajectory of causality explained and calculate the complementary metrics such as the polyfactorial index and causality gain as the difference between the causality explained by the first exposure and the maximum causality explained. Assuredly, even if the maximum causality explained remains similar, the causality explained trajectory as we have defined it, as well as the polyfactorial index and the causality gain cannot be interpreted. As an illustration, we have run the causality explained model on the 13 diseases where PheCodes are ordered with increasing levels of individual causality explained (Supplementary Figure 14).
